# Prognostic and predictive biomarkers in patients with COVID-19 treated with tocilizumab in a randomised controlled trial

**DOI:** 10.1101/2020.12.23.20247379

**Authors:** Jennifer Tom, Min Bao, Larry Tsai, Aditi Qamra, David Summers, Montserrat Carrasco-Triguero, Jacqueline McBride, Carrie M Rosenberger, Celia J F Lin, William Stubbings, Kevin G Blyth, Jordi Carratalà, Bruno François, Thomas Benfield, Derrick Haslem, Paolo Bonfanti, Cor H van der Leest, Nidhi Rohatgi, Lothar Wiese, Charles Edouard Luyt, Farrah Kheradmand, Ivan O Rosas, Fang Cai

## Abstract

**Background:** Retrospective observational studies suggest that interleukin-6 (IL-6), C-reactive protein (CRP), lactate dehydrogenase (LDH), ferritin, lymphocytes, monocytes, neutrophils, D-dimer, and platelets are associated with disease progression, treatment outcomes, or both, in patients with COVID-19 pneumonia. We explored these candidate prognostic and predictive biomarkers with efficacy outcomes after treatment with tocilizumab, an anti–IL-6 receptor antibody using data from the COVACTA trial for patients hospitalised with severe COVID-19 pneumonia.

**Methods:** Candidate biomarkers were measured in 295 patients in the tocilizumab arm and 142 patients in the placebo arm. Efficacy outcomes assessed were clinical status on a seven-category ordinal scale (1, discharge; 7, death), mortality, time to hospital discharge, and mechanical ventilation (if not receiving it at randomisation) through day 28. Prognostic and predictive biomarkers were evaluated continuously with proportional odds, binomial or Fine-Gray models, and additional sensitivity analyses.

**Findings:** Modelling in the placebo arm showed all candidate biomarkers except LDH and D-dimer were strongly prognostic for day 28 clinical outcomes of mortality, mechanical ventilation, clinical status, and time to hospital discharge. Modelling in the tocilizumab arm showed a predictive value of ferritin for day 28 clinical outcomes of mortality (predictive interaction p=0.03), mechanical ventilation (predictive interaction p=0.01), and clinical status (predictive interaction p=0.02) compared with placebo.

**Interpretation:** Multiple biomarkers prognostic for clinical outcomes were confirmed in COVACTA. Ferritin was identified as a predictive biomarker for the effects of tocilizumab in the COVACTA patient population; high ferritin levels were associated with better clinical outcomes for tocilizumab compared with placebo at day 28.

**RESEARCH IN CONTEXT:** 

**Evidence before this study:** The efficacy and safety of the anti–interleukin-6 receptor antibody tocilizumab in the treatment of patients hospitalised with COVID-19 pneumonia was investigated in COVACTA, a double-blind, randomised, placebo-controlled trial. The primary endpoint of improved clinical status on a seven-category ordinal scale (1, discharged/ready for discharge; 7, death) at day 28 was not met in this trial. Among the secondary endpoints, no difference in mortality at day 28 was observed, but time to hospital discharge was shorter in the tocilizumab group. Subgroup analysis suggested there might be a treatment benefit in patients grouped according to their ordinal scale category at baseline.

We searched PubMed on September 14, 2020, using the terms “tocilizumab AND (COVID-19 OR coronavirus) AND biomarker” with no language or date restrictions. The search retrieved 18 articles, four of which identified laboratory measures as potential biomarkers in patients who received tocilizumab for the treatment of COVID-19 pneumonia. The biomarkers reported in these studies include interleukin-6, C-reactive protein, ferritin, fibrinogen, liver transaminases, lymphocytes, platelets, and D-dimer. However, these previous studies were single-centre, retrospective, observational studies. Larger, prospective, controlled trials are needed to investigate potential prognostic and predictive biomarkers to assess the outcomes and response to treatments for COVID-19.

**Added value of this study:** This exploratory analysis of data from COVACTA demonstrated interleukin-6, C-reactive protein, ferritin, neutrophils (percentage and absolute count), neutrophil-to-lymphocyte ratio, lymphocytes (percentage and absolute count), monocytes (percentage), and platelets as strong prognostic biomarkers in patients hospitalised with severe COVID-19 pneumonia. More important, ferritin showed predictive value for tocilizumab treatment effects on day 28 clinical outcomes of mortality, mechanical ventilation (among the subgroup of patients not receiving mechanical ventilation at randomisation), and clinical status compared with placebo.

**Implications of all the available evidence:** In patients with elevated levels of ferritin at baseline, tocilizumab decreased the probability of death, mechanical ventilation, and worsening clinical status at day 28 compared with placebo, suggesting that ferritin might be useful as a predictive biomarker of efficacy outcomes for tocilizumab in patients with severe COVID-19 pneumonia.

## Introduction

Coronavirus disease-19 (COVID-19) elicits an initial innate antiviral pro-inflammatory immune response and a second phase that reflects an adaptive immune response.^1^ Pro-inflammatory cytokines, particularly interleukin-6 (IL-6) and tumour necrosis factor-α, may be involved in the immune dysregulation reported in some patients with COVID-19, possibly resulting in more severe clinical manifestations.^2,3^ Elevated levels of IL-6 are associated with severe COVID-19,^4^ and preliminary findings from case-control and retrospective cohort studies^5,6^ support investigation of tocilizumab, an anti–IL-6 receptor alpha antibody, to treat severe COVID-19 pneumonia.

COVACTA was a global, double-blind, randomised, placebo-controlled, phase 3 trial of tocilizumab in patients hospitalised with severe COVID-19 pneumonia. The primary endpoint of improved clinical status on a seven-category ordinal scale at day 28 was not met. Among secondary endpoints, no difference was observed in mortality at day 28; however, time to hospital discharge was shorter in the tocilizumab group. Subgroup analysis suggested there might be a reduced risk for intensive care unit (ICU) transfer among patients not in the ICU at randomisation and a potential reduction in clinical failure (death, mechanical ventilation, ICU transfer, or study withdrawal) among patients not receiving mechanical ventilation at randomisation. Because the primary endpoint of the study was not met, additional studies are ongoing to investigate these findings further.^7^

Patients with COVID-19 have diverse symptoms and complications, from pneumonia to acute respiratory distress syndrome, multiorgan failure, and death despite receiving current standard care.^8^ The heterogeneity in clinical presentation and the pathological manifestations of COVID-19 present major challenges in investigating effective treatments and identifying optimal biomarkers that might help prognosticate or predict response to treatments. Laboratory measures of hyperinflammation (IL-6, C-reactive protein [CRP]), macrophage activation (ferritin), dysregulated immune cells (lymphocytes, monocytes, neutrophils), tissue damage (lactate dehydrogenase [LDH]), and coagulopathy (D-dimer, platelets) have been reported as risk factors for poor prognosis in COVID-19,^4,9-13^ and most of them are mechanistically linked to IL-6 activity.^14-16^ This suggests they might be useful not only as prognostic but also as predictive biomarkers of the effects of IL-6 pathway blockade. To date, these biomarkers have been investigated only in retrospective observational studies of COVID-19.

We aimed to determine the prognostic and predictive value of previously identified candidate biomarkers regarding clinical outcomes in a pre-specified exploratory analysis of the COVACTA trial.

## Methods

### Patients and study design

COVACTA (ClinicalTrials.gov, NCT04320615) was a randomised, placebo-controlled, double-blind, global, multicentre, phase 3 trial investigating the efficacy and safety of tocilizumab versus placebo in patients hospitalised with severe COVID-19 pneumonia receiving standard care according to local practice.^7^ Eligible adult patients—stratified by geographic region (North America, Europe) and mechanical ventilation (yes, no)—were to receive intravenous tocilizumab or placebo in a 2:1 ratio. Key inclusion criteria were hospitalisation with SARS-CoV-2 infection confirmed by polymerase chain reaction of any specimen and blood oxygen saturation (SpO_2_) ≤93% or partial pressure of oxygen/fraction of inspired oxygen (PaO_2_/FiO_2_) <300 mm/Hg despite receiving local standard care (which could include antivirals or low-dose steroids). Key exclusion criteria were active or suspected infection (other than SARS-CoV-2); alanine aminotransferase or aspartate aminotransferase >10× the upper limit of normal; absolute neutrophil count <1000/mL; and platelet count <50,000/mL. The primary endpoint of the trial was clinical status assessed using a seven-category ordinal scale. Secondary endpoints included mortality at day 28, incidence of mechanical ventilation (in patients not receiving mechanical ventilation at randomisation), and time to hospital discharge. Informed consent was obtained for all enrolled patients. The study was conducted in accordance with the International Council for Harmonization E6 guideline for good clinical practice and the Declaration of Helsinki or local regulations, whichever afforded greater patient protection. The protocol was reviewed and approved by all appropriate institutional review boards and ethics committees (see Appendix 1 for the complete list).

### Biomarker analysis

Biomarkers in peripheral blood were assessed at baseline to examine their potential prognostic value and potential predictive value for tocilizumab efficacy. Efficacy was determined in relation to the clinical status assessed on the seven-category ordinal scale at day 28 (primary), mortality at day 28, time to hospital discharge, and requirement for mechanical ventilation by day 28 (in patients not receiving it at randomisation). These four endpoints were selected to ensure the signal was robust and consistent across endpoints. Imputation rules for efficacy endpoints were assigned as published.^7^ Pre-specified candidate biomarkers included IL-6 and CRP as markers of hyperinflammation, ferritin as an acute-phase protein and as a marker of macrophage activation, LDH as a marker of tissue damage, lymphocytes as a marker of dysregulated immune response, and D-dimer as a marker of coagulopathy. The potential for neutrophil, monocyte, and platelet counts as biomarkers was also explored. IL-6 levels were measured using an immunoassay method validated at QPS (Quantikine ELISA; R&D Systems Minneapolis, MN). CRP levels were measured using an in vitro diagnostic method validated at PPD (Roche Cobas; Roche Diagnostics, Indianapolis, IN). All other biomarkers were assessed using standard clinical chemistry and haematology methods available at the local clinical laboratories.

### Statistical analysis

Biomarkers were assessed in the modified intention-to-treat population (any randomly assigned patients who received study medication). Subgroup analysis was conducted in patients in ordinal scale categories 4 (ICU or non-ICU hospital ward, requiring non-invasive ventilation or high-flow oxygen) and 5 (ICU, requiring intubation and mechanical ventilation) at baseline (i.e. patients who might benefit from tocilizumab).^7^ Histograms, scatterplots, and tables were generated to assess the completeness of data by treatment arm at baseline, determine the balance of biomarker levels at baseline, and identify outliers. Biomarkers not normally distributed were log transformed before analysis, among them D-dimer, ferritin, IL-6, lymphocytes (absolute), neutrophil-to-lymphocyte ratio, and white blood cells. Pearson correlation coefficient was calculated between outcomes and biomarkers, between the baseline covariates and biomarkers, and between individual biomarkers. Six candidate biomarkers assessed (IL-6, CRP, ferritin, LDH, lymphocytes, D-dimer) were pre-specified in a biomarker analysis plan. No adjustments were made for multiple comparisons.

Prognostic modelling was assessed in the placebo arm only, controlling for the following covariates: mechanical ventilation status at randomisation (yes/no), antiviral use (yes/no), steroid use (yes/no), age, sex, and region (Europe/North America). None of the endpoints were considered primary because the motivation was to identify a robust, consistent signal across endpoints. For each endpoint, sensitivity analyses were performed on the placebo arm only using unadjusted analysis, on the tocilizumab and placebo arms adjusting for the same covariates plus treatment arm, and on the tocilizumab and placebo arms controlling only for treatment arm. Candidate prognostic biomarkers were assessed using a proportional odds model with the ordinal scale score at day 28 as a dependent variable and biomarker and covariates (depending on the model) as independent variables. Odds ratios, confidence intervals, and p-values were reported, and proportional odds assumptions were assessed graphically. A Fine-Gray model was fit for time to discharge, with death as a competing risk. A Cox proportional hazards model was fit as a sensitivity analysis. A binomial model with the outcome as a dependent variable, biomarkers as independent variables, and covariates (depending on the model) was used for binary outcomes (death, discharge, mechanical ventilation).

Candidate predictive biomarkers were modelled in the same way as the candidate prognostic biomarkers, with the addition of an interaction term between continuous biomarker value and treatment. All reported predictive p-values rely on the interaction p-value term except for those generated in the multivariate model assessing multiple biomarkers. Additionally, tertile analysis of predictive biomarkers was performed by creating vectors for each tertile (low, medium, high) and fitting a single model with interaction terms for medium and high tertiles with treatment. Treatment effects within each tertile were then calculated from the estimates. No cut-point optimisation was performed, but analysis and visualisation were performed using tertiles and quartiles. Combined predictive biomarkers were assessed by dichotomising the biomarkers using median values as cut-offs.

To support the predictive findings, an additional analysis was conducted using data from the placebo arm of COVACTA and data from a phase 2 trial of tocilizumab in moderate to severe COVID-19 pneumonia (ClinicalTrials.gov NCT04363736; MARIPOSA) (Appendix Table S1). Data from the tocilizumab 8-mg/kg arm of MARIPOSA were analysed and compared with data from the placebo arm of COVACTA, followed by sensitivity analysis including pooled data from the tocilizumab 8-mg/kg and 4-mg/kg arms of MARIPOSA. Inclusion criteria were matched, and the subset of patients with severe disease in MARIPOSA (similar to the COVACTA population) was included but patients with moderate disease were excluded. Propensity scores were calculated based on the following covariates, which could impact treatment assignment or outcome: ordinal baseline score (captures mechanical ventilation status), age, sex, antiviral use (yes/no), and corticosteroid use (yes/no). Overlapping support was checked for the propensity score distributions. Matching was performed using the MatchIt (version 3.0.2) algorithm, which matched the treatment groups from MARIPOSA and the control group from COVACTA using propensity scores with the nearest neighbour algorithm. Weighting was performed using inverse probability weighting according to the ATT estimand.^17^ Success of weighting and matching was assessed using Love plots, which chart the standardised differences of variables (standardised mean difference [SMD] = *x*_treatment – *x*_control)/(pooled standard deviation), where SMD <0.1 or SMD <0.25 was considered acceptable,^18,19^ and histograms (for categorical variables) or density plots (for continuous variables) before and after weighting were used for visual comparison. Several methods were used to estimate the treatment difference for subgroup analysis and the interaction term between the continuous predictive biomarker and treatment: ATT estimand via propensity score weighting (primary analysis method); propensity score regression; naive; propensity score matching.

## Results

### Biomarker levels at baseline

Baseline biomarker levels (Appendix Figure 1A), except IL-6 and CRP, were generally balanced between the treatment arms in COVACTA (Table 1; Appendix Figure 1B). Median levels were higher in the tocilizumab than the placebo arm for IL-6 (88.1 vs 70.3 ng/L) and CRP (169.3 vs 151.9). Median baseline levels of IL-6, CRP, ferritin, D-dimer, LDH, and neutrophils (absolute value and percentage) were above normal ranges, and lymphocytes (absolute value and percentage) were below normal ranges (neutrophil-to-lymphocyte ratio was elevated). Monocytes (percentages), platelet counts, and total leukocyte counts were within normal ranges. Limited correlation was observed between the pre-specified biomarkers (Appendix Figure 1C).

**Table 1:** Biomarker levels at baseline (modified intention-to-treat population)

| **Biomarker** | **Normal range** | **Placebo** | **Tocilizumab** |
| --- | --- | --- | --- |
| IL-6, ng/mL* | $\leq 0.007$ | n=99 | n=233 |
| Mean (SD) |  | 192.2 (368.7) | 201.9 (418.4) |
| Median (range) |  | 70.3 (3.1–2810) | 88.1 (3.1–4020) |
| CRP, mg/L | $\leq 5$ | n=126 | n=256 |
| Mean (SD) |  | 177.8 (117.1) | 187.8 (119.8) |
| Median (range) |  | 151.9 (1.6–500) | 169.3 (1.1–500) |
| Ferritin, pmol/L* |  | n=124 | n=240 |
| Mean (SD) | 27–$\leq 337$ (women) | 3792 (7463) | 3069 (3113) |
| Median (range) | 27–$\leq 674$ (men) | 2168 (96.9–75300) | 2250 (3.6–24045) |
| D-dimer, $\mu\text{g/mL FEU}^*$ | $\leq 0.5$ | n=66 | n=131 |
| Mean (SD) |  | 4.2 (7.6) | 4.6 (8.4) |
| Median (range) |  | 1.2 (0.3–46.7) | 1.3 (0.2–58.1) |
| LDH, IU/L | 105–333 | n=121 | n=243 |
| Mean (SD) |  | 469.7 (291.7) | 479.4 (303.5) |
| Median (range) |  | 422 (1.3–2323) | 430 (0.7–3282) |
| Leukocytes $10^9/\text{L}^*$ | 4.5–11 | n=140 | n=280 |
| Mean (SD) |  | 9.2 (4.1) | 9.3 (4.5) |
| Median (range) |  | 8.5 (2.4–22.4) | 8.3 (2.7–28.2) |
| Lymphocytes $10^9/\text{L}^*$ | 0.9–2.9 | n=139 | n=288 |
| Mean (SD) |  | 0.96 (0.83) | 0.98 (0.57) |
| Median (range) |  | 0.9 (0–8.9) | 0.9 (0–5.4) |
| Lymphocytes, % | 20–40 | n=133 | n=268 |
| Mean (SD) |  | 13 (10) | 12 (7) |
| Median (range) |  | 11 (0–55) | 11 (0–48) |
| Neutrophils 10<sup>9</sup>/L | 1·7–7 | n=139 | n=291 |
| Mean (SD) |  | 7·5 (3·9) | 7·6 (4·1) |
| Median (range) |  | 7·2 (0·9–23·1) | 6·8 (1·0–24·6) |
| Neutrophils, % | 40–6 | n=134 | n=268 |
| Mean (SD) |  | 79 (12) | 79 (9) |
| Median (range) |  | 82 (24–98) | 81 (44–99) |
| Neutrophils/lymphocytes, ratio* | 1–3 | n=132 | n=265 |
| Mean (SD) |  | 65·0 (566·5) | 64·8 (683·6) |
| Median (range) |  | 7·1 (0·4–6509) | 7·5 (1·2–10463) |
| Monocytes, % | 2–8 | n=131 | n=269 |
| Mean (SD) |  | 6 (4) | 6 (4) |
| Median (range) |  | 5 (0–19) | 5 (0–32) |
| Platelets, 10<sup>9</sup>/L | 150–400 | n=142 | n=295 |
| Mean (SD) |  | 262·0 (117·7) | 265·7 (113·3) |
| Median (range) |  | 240 (53–814) | 253 (10·2–825) |

\*Log transformed for all additional analyses.
CRP=C-reactive protein; FEU=fibrinogen equivalent units; IL-6=interleukin-6; LDH=lactate dehydrogenase.

**Figure 1:**
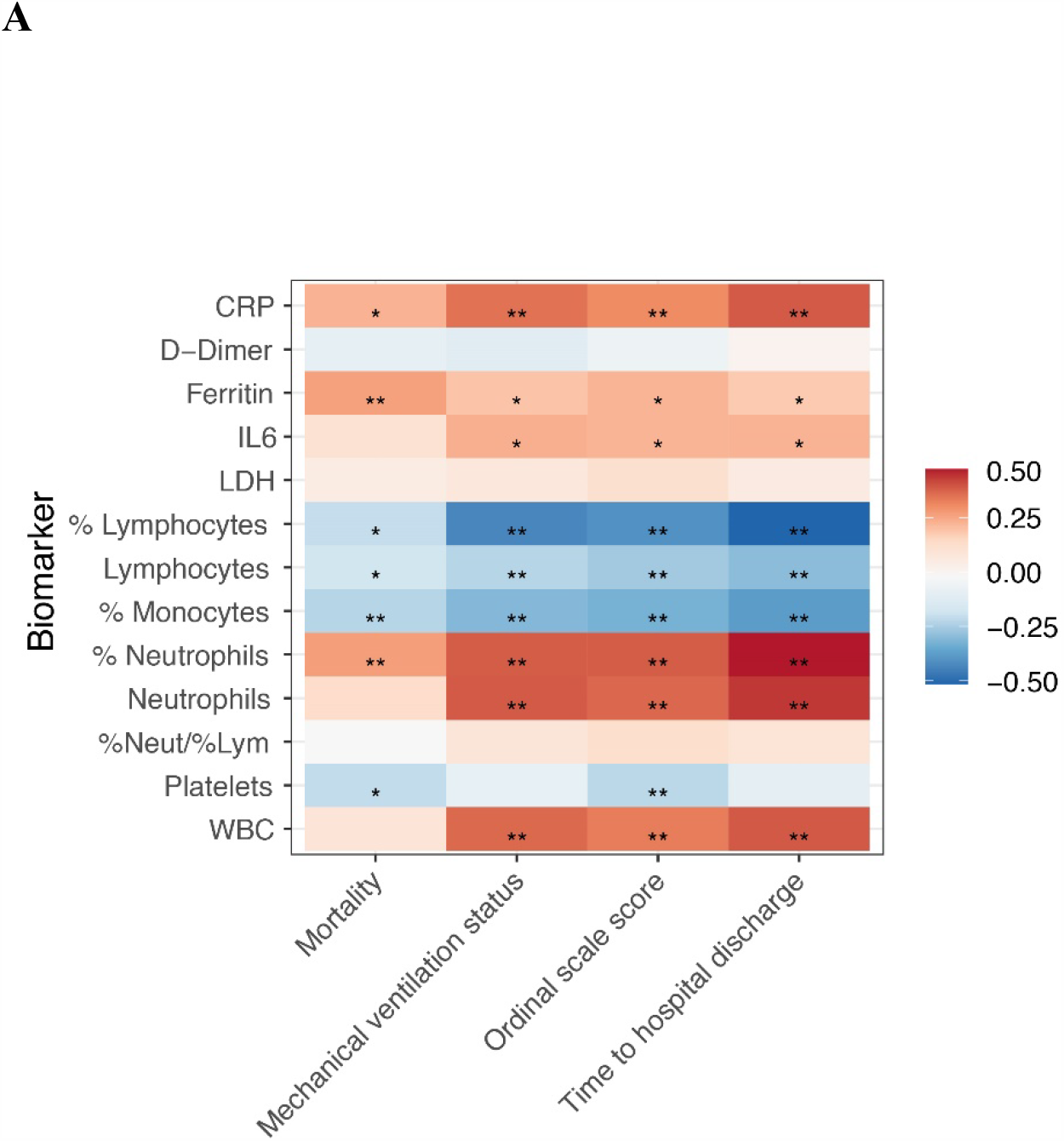
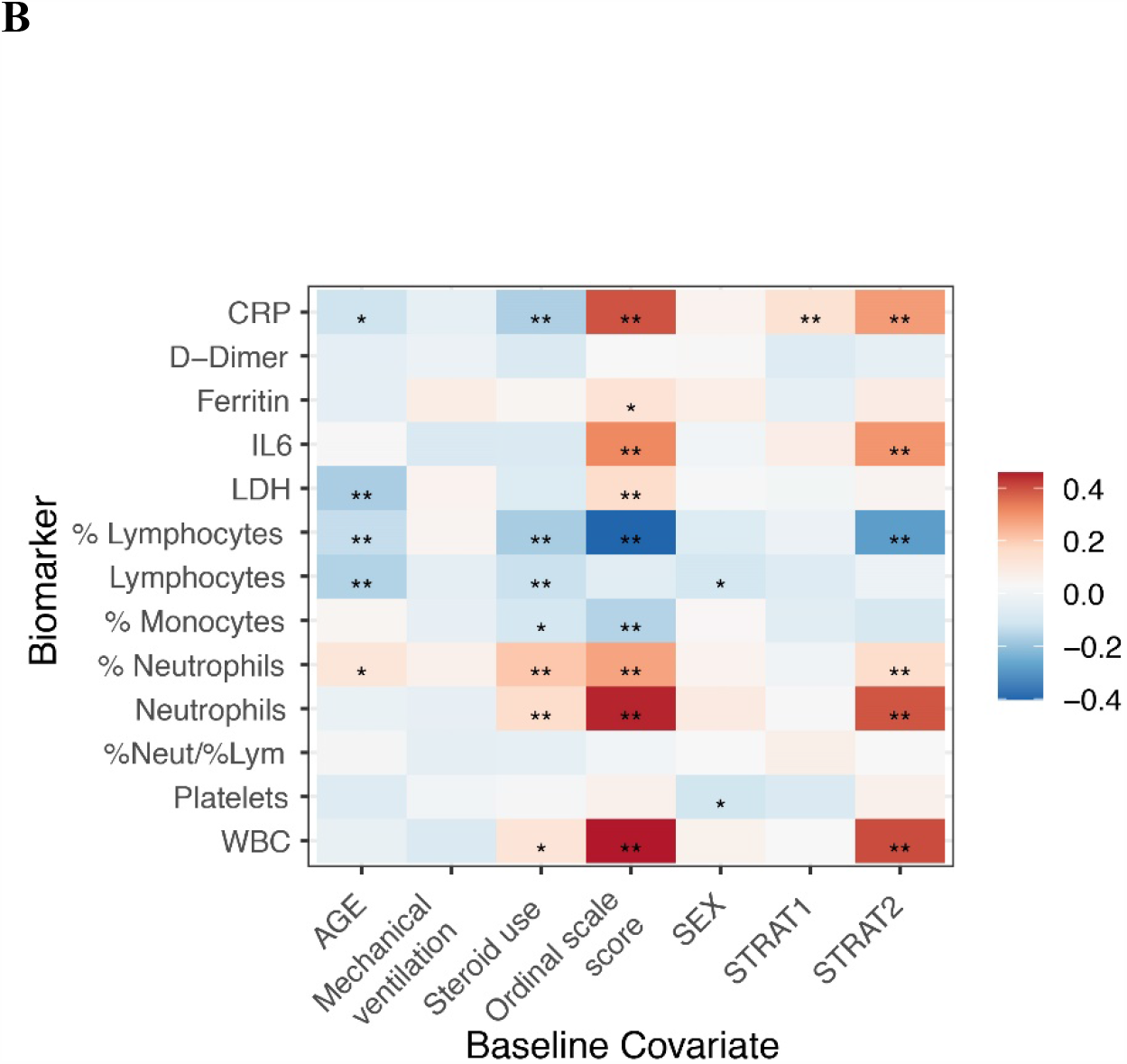
Correlation between (A) baseline biomarkers and clinical outcomes and (B) biomarkers and baseline covariates. p values are based on Pearson correlation unadjusted for covariates, placebo arm only (n=142). *p≤0·05; **p≤0·01. CRP=C-reactive protein; IL-6=interleukin-6; LDH=lactate dehydrogenase; PBO=placebo; TCZ=tocilizumab; WBC=white blood cell.

### Evaluation of prognostic biomarkers

All biomarkers examined, with the exception of LDH, D-dimer, and neutrophil-to-lymphocyte ratio, showed a correlation with clinical outcomes, including mortality, mechanical ventilation, ordinal scale score, and time to hospital discharge, at day 28 (Figure 1A). Some biomarkers showed correlation with baseline covariates; for example, IL-6, CRP, neutrophils (absolute value and percentage), and total leukocyte count were positively correlated with higher baseline ordinal scale category and mechanical ventilation at randomisation (yes, no) (Figure 1B). Adjusting for covariates, all biomarkers except LDH and D-dimer showed robust evidence across different sensitivity analyses, supporting their prognostic value for clinical outcomes (Appendix Figure S2). Prognostic modelling showed a consistent direction of effect across all clinical outcomes at day 28 for mortality, mechanical ventilation, ordinal scale score, and time to hospital discharge (Figure 2A; Appendix Figure S2). Analysis of the ordinal scale at day 28 according to baseline continuous ferritin revealed a potential prognostic effect of ferritin in the placebo arm (p=0.01; n=124).

**Figure 2:**
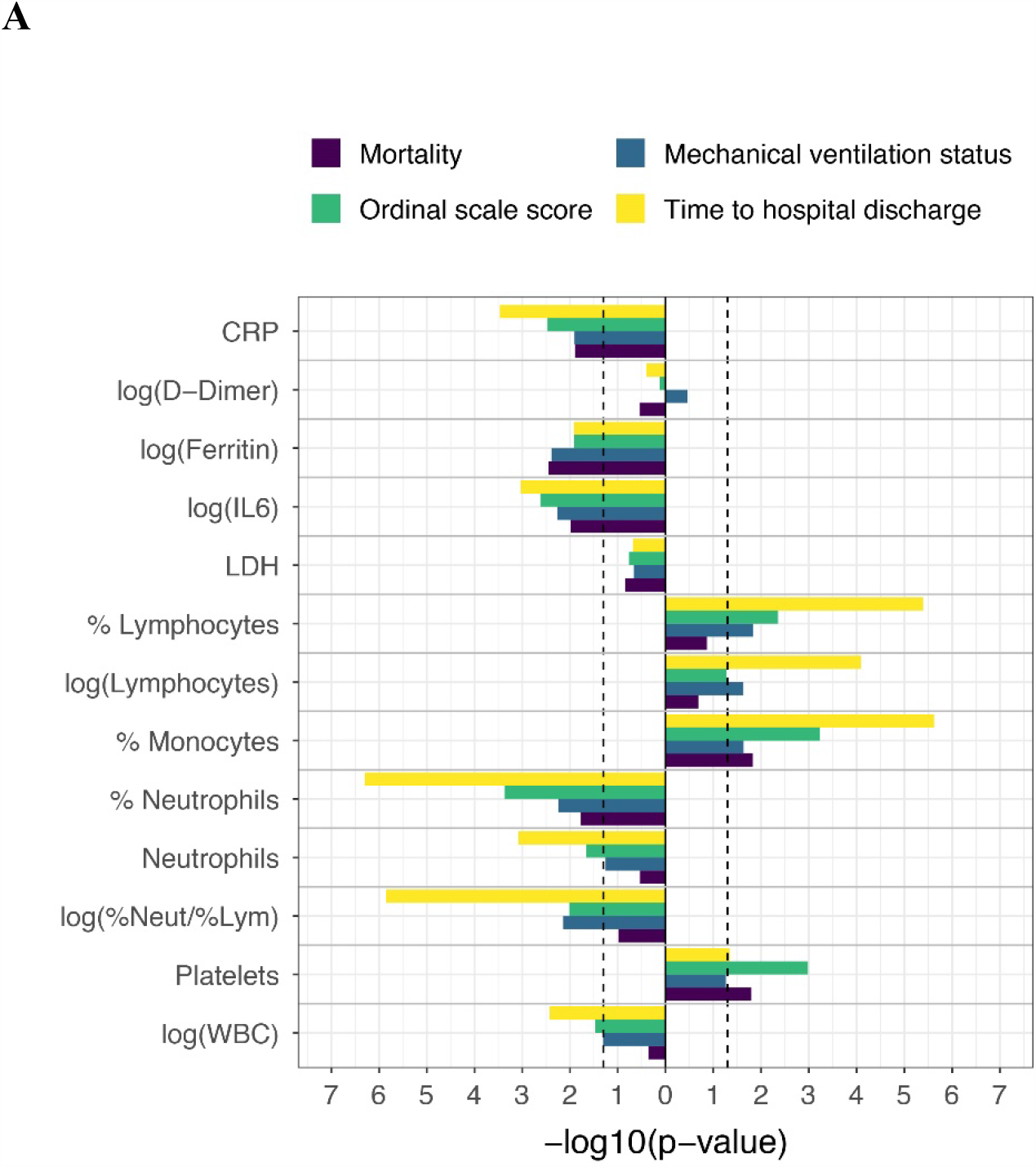
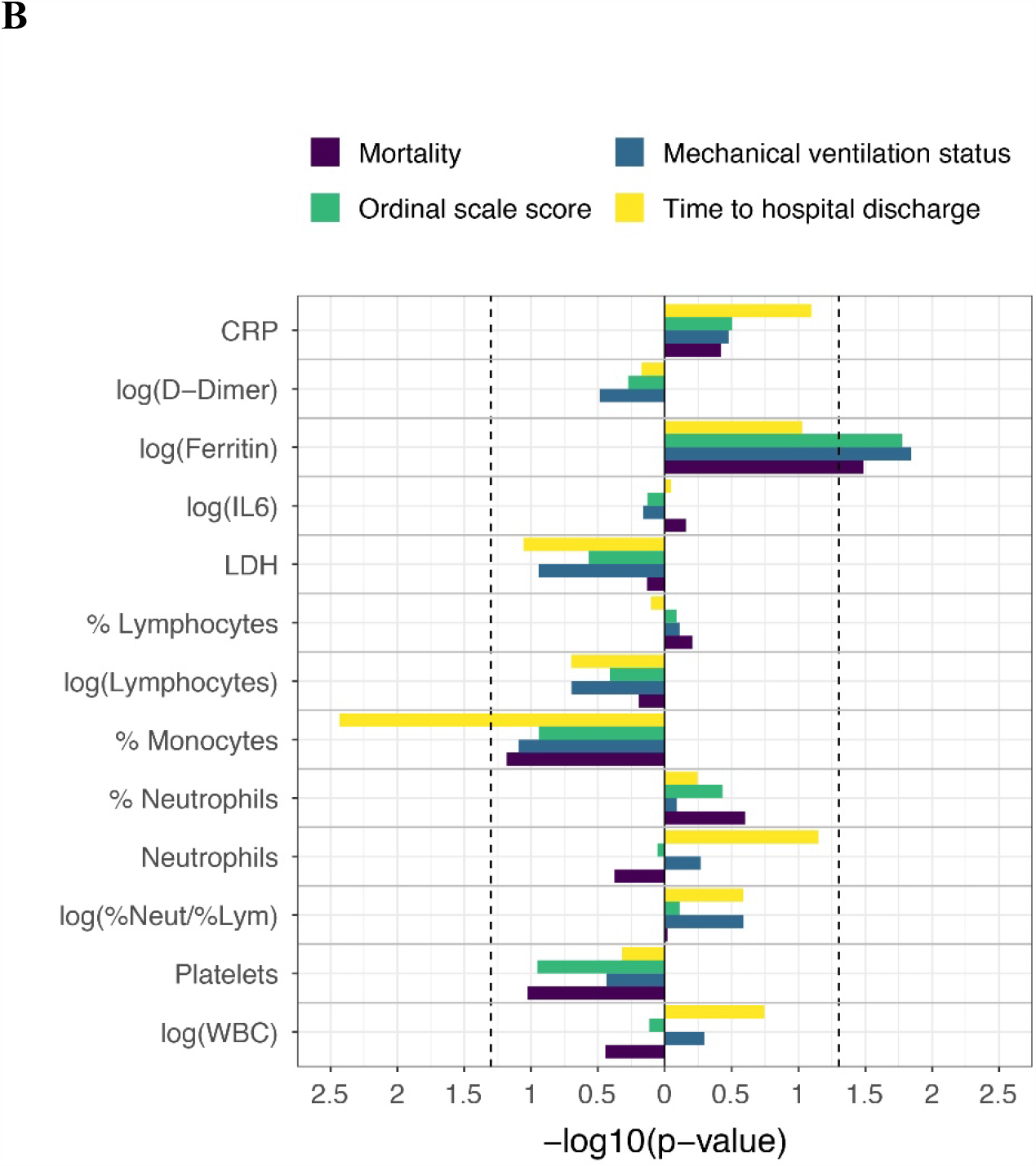
Efficacy outcomes. (A) Placebo-adjusted, scaled prognostic model and (B) predictive model. Model adjusted for the stratification factors of region (Europe, North America) and mechanical ventilation (yes, no) as well as age, sex, baseline antiviral use (yes, no), and baseline steroid use (yes, no). Vertical dashed lines indicate p=0·05. Outcomes are at day 28 and have been aligned so that bars pointing in the same direction have the same direction of effect. Last observation carried forward was used for ordinal scale score at day 28. CRP=C-reactive protein; LDH=lactate dehydrogenase; PBO=placebo; WBC=white blood cell.

### Evaluation of predictive biomarkers

Predictive modelling for all assessed biomarkers except IL-6, lymphocytes (percentage), neutrophils (absolute), and leucocytes showed a consistent direction of effect across all clinical outcomes (Figure 2B). Ferritin was identified as a predictive biomarker for the effects of tocilizumab in COVACTA (Appendix Figure S3). No other biomarkers had predictive value for the effects of tocilizumab except monocytes (percentage), with low values predictive for time to hospital discharge (Appendix Figure S3). Analysis of the ordinal scale at day 28 according to continuous baseline ferritin values showed that placebo patients with higher baseline levels had worse clinical status, whereas clinical outcomes at day 28 were similar across baseline ferritin tertiles in tocilizumab patients (n=364; predictive effect interaction, p=0.02; visualised using tertiles in Figure 3A). Modelled data showed a predictive effect for ferritin as a biomarker for tocilizumab effects on mortality by day 28 (predictive interaction, p=0.03) (Figure 3B) and mechanical ventilation by day 28 (predictive interaction, p=0.01) (Figure 3C). Tocilizumab treatment decreased the probability of mechanical ventilation and mortality by day 28 compared with placebo in patients with elevated levels of ferritin based on continuous assessment across a range of ferritin levels. Although time to hospital discharge was nominally significantly shorter with tocilizumab than placebo in COVACTA,^7^ ferritin was not a significant predictive biomarker for the effect of tocilizumab on time to hospital discharge (p=0.09) (Figure 3D); however, the direction of effect was consistent with that observed for the ordinal scale category and mortality outcomes.

**Figure 3:**
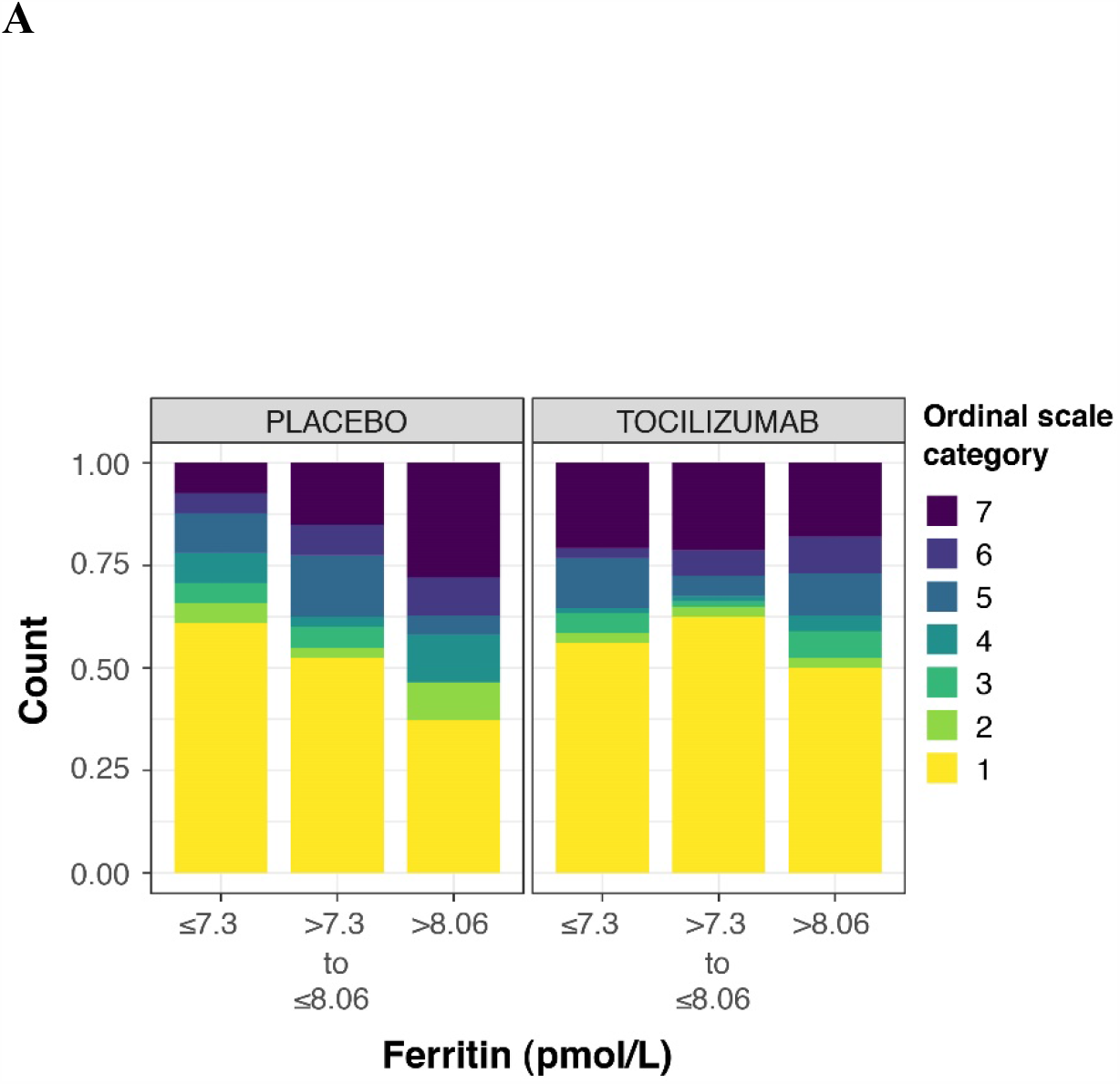
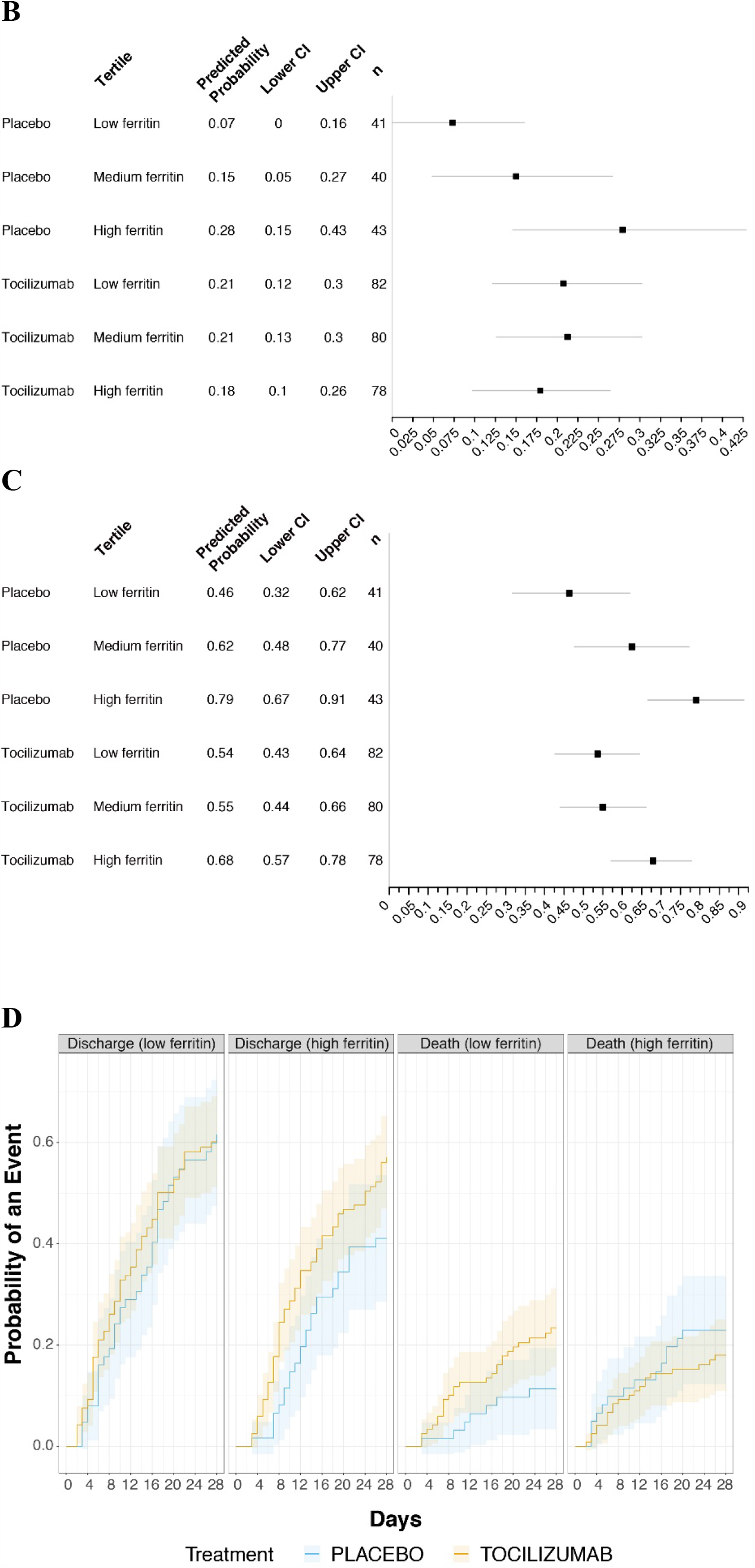
Ferritin as a predictor for efficacy outcomes. (A) Day 28 all comers, (B) unadjusted tertiles model-predicted probability of death to day 28, (C) unadjusted tertiles model-predicted probability of mechanical ventilation to day 28 (in patients who did not receive mechanical ventilation at randomisation), and (D) cumulative incidence of hospital discharge and death to day 28. Seven-category ordinal scale: 1, discharged or ready for discharge; 2, non–ICU hospital ward, not requiring supplemental oxygen; 3, non–ICU hospital ward, requiring supplemental oxygen; 4, ICU or non–ICU hospital ward, requiring non-invasive ventilation or high-flow oxygen; 5, ICU, requiring intubation and mechanical ventilation; 6, ICU, requiring extracorporeal membrane oxygenation or mechanical ventilation and additional organ support; 7, death. Ferritin values were log transformed (sample size, n=364). Ferritin tertile cut-off values were 3·63 pmol/L (minimum), 1480·77 pmol/L, 3150·29 pmol/L, and 75299·67 pmol/L (maximum) in an unadjusted model showing predicted probability and 95% CI (B, C). Cumulative incidence was determined by Aalen-Johansen estimator, with an arbitrary median cut-point of 7·7 on logarithmic scale (D). ICU=intensive care unit.

Additional evidence of the value of ferritin as a predictive biomarker of the effects of tocilizumab was generated using unpublished data from the phase 2 MARIPOSA trial of tocilizumab in COVID-19. The binary outcomes of mortality and mechanical ventilation were assessed. Analysis comparing data from patients with severe disease in the tocilizumab 8-mg/kg arm from MARIPOSA with data from the placebo arm of COVACTA supported the finding that ferritin has predictive value as a biomarker for mortality (ATT estimand based, p=0.044); a similar pattern was observed for mechanical ventilation (Appendix Figure S4).

Analysis of the ordinal scale at day 28 and mortality at day 28 suggested that baseline IL-6 is not a predictive marker of tocilizumab efficacy (Appendix Figure S5). Analysis of combinations of the biomarkers ferritin, IL-6, and CRP (n=257) as well as ferritin, LDH, and D-dimer (n=165) did not show any synergistic effect between biomarkers (Appendix Table S2), further suggesting that ferritin alone was the strongest predictive biomarker for the effects of tocilizumab in COVACTA.

### Subgroup analyses

The subgroup of patients who required high-flow oxygen or non-invasive or invasive mechanical ventilation without other advanced life support (ordinal scale categories 4 and 5) at baseline had higher baseline ferritin levels than patients in all other ordinal scale categories (Appendix Figure S6, Table S3), and those in the tocilizumab arm had better clinical outcomes than those in the placebo arm. Analysis of this subgroup using a continuous scale for baseline ferritin levels also demonstrated a predictive association of baseline ferritin levels and the effects of tocilizumab for mortality (interaction p=0.02). For clinical utility, tertile analysis was also performed (Figure 4, Appendix Figure S7). The predictive signal of ferritin, treated as a continuous biomarker, was also significant for clinical status by ordinal scale at day 28 (interaction p=0.01), as was incidence of mechanical ventilation by day 28 in patients who did not receive mechanical ventilation at randomisation (interaction p=0.01).

**Figure 4:**
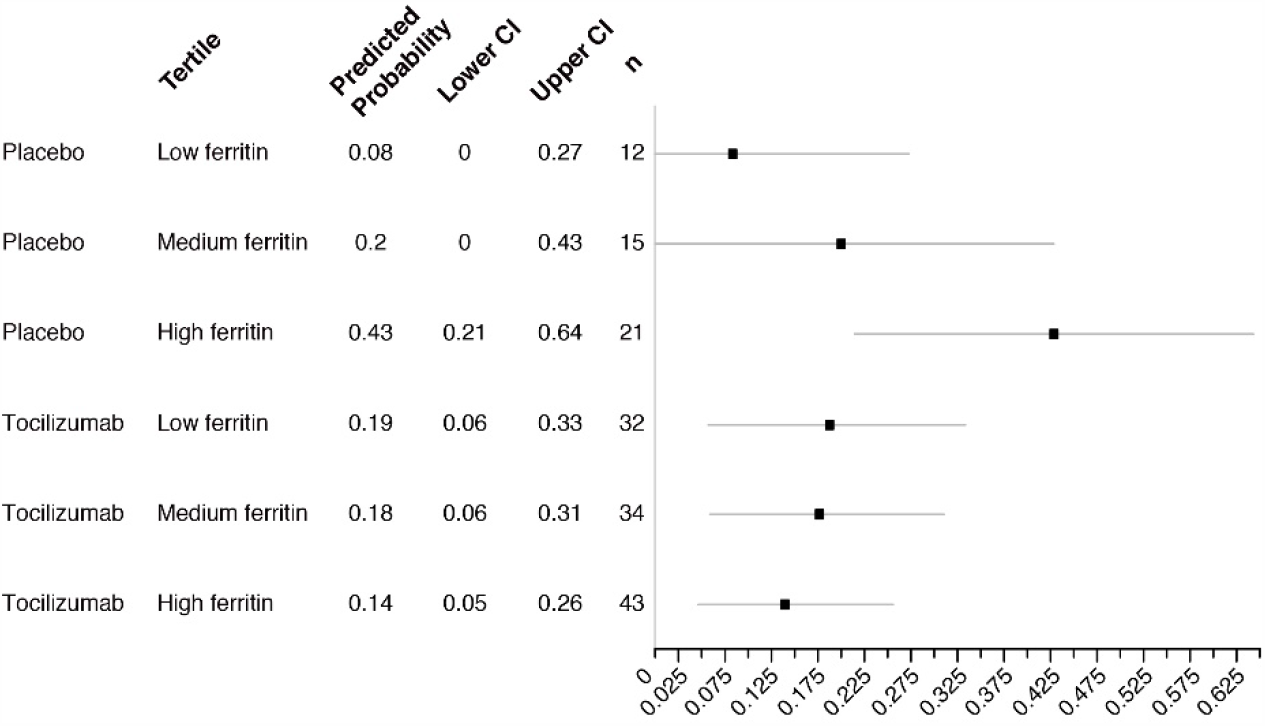
Predictive biomarker assessment for death by day 28 according to baseline ferritin levels. Subset of patients in ordinal scale categories 4 and 5 at baseline in COVACTA; n=157. Ferritin values were log transformed. Shown are predicted probabilities for death within each tertile of ferritin (tertile cut-off values were 3·63 pmol/L [minimum], 1480·77 pmol/L, 3150·29 pmol/L, and 75299·67 pmol/L [maximum]) showing predicted probability and 95% CI based on an unadjusted model (calculated based on all-comers) fit using samples restricted to those for patients in ordinal scale categories 4 and 5 at baseline.

Baseline IL-6 levels were predictive of the effect of tocilizumab in this subgroup for time to hospital discharge only (Figure 5, Appendix Figure S8) (adjusted Fine-Gray p=0.03), but this was not statistically significant in the sensitivity analysis (Cox proportional hazards adjusted p=0.09). Furthermore, baseline IL-6 levels were not predictive for tocilizumab effects on hospital discharge (yes vs no; p=0.13) or any other outcome in this subgroup.

**Figure 5:**
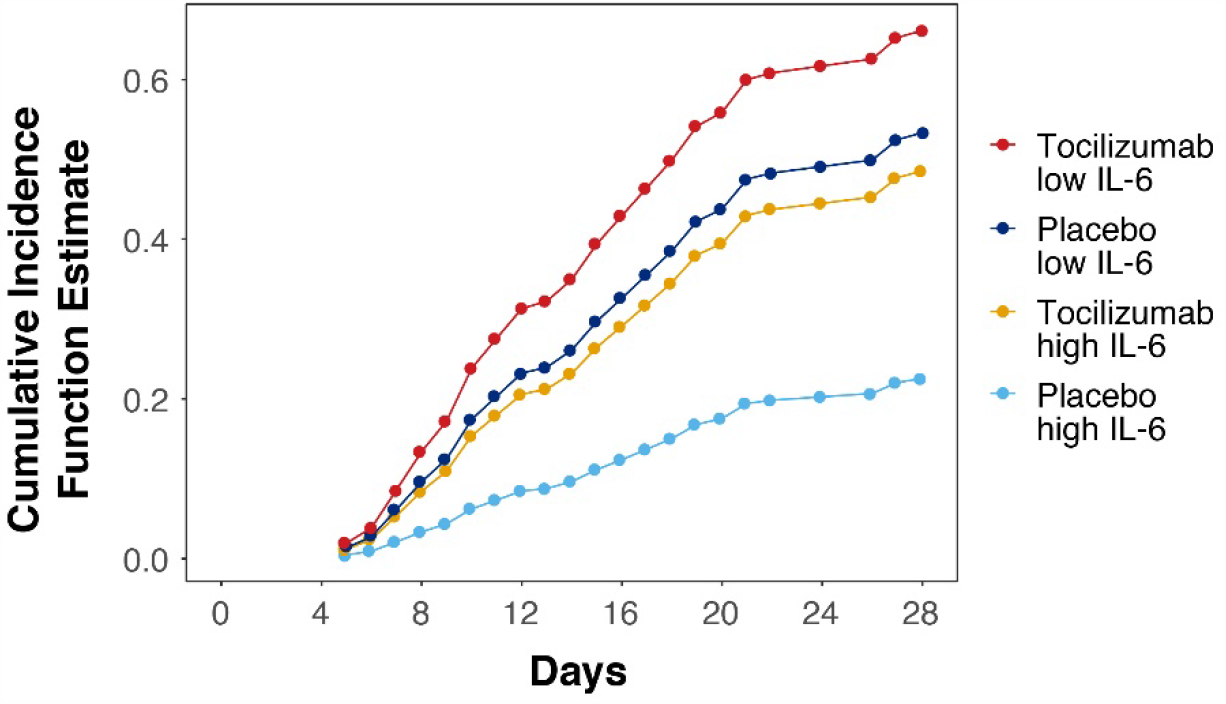
Time to hospital discharge by baseline IL-6 levels. (subset of patients in ordinal scale categories 4 and 5 at baseline in COVACTA; n=157). Ferritin values were log transformed. Figure shows the cumulative incidence function for time to hospital discharge based on the median IL-6 cut-off value. IL-6=interleukin-6.

## Discussion

Our exploratory analysis of the COVACTA trial is the first, to our knowledge, to report on prognostic and predictive biomarkers from a global, multicentre, randomised controlled trial in patients hospitalised with COVID-19 pneumonia. IL-6, CRP, ferritin, LDH, lymphocytes, neutrophils, monocytes, D-dimer, and platelets are among the biomarkers identified in case series and observational studies as potential prognostic biomarkers for disease outcomes in COVID-19.^9,12,20^

IL-6, CRP, ferritin, LDH, lymphocyte, neutrophil, and D-dimer values were abnormal at baseline in the COVACTA study population, consistent with the pathological manifestations of hyperinflammation, macrophage activation, tissue damage, dysregulated immune cells, and coagulopathy commonly observed in patients with severe COVID-19.^4,9-13^ Only a modest correlation was observed between the biomarkers representing different manifestations, potentially reflecting the overlapping yet differentiated pathogenic pathways in COVID-19. Elevated levels of IL-6, CRP, ferritin, and neutrophils and decreased levels of lymphocytes demonstrated robust and consistent prognostic value as biomarkers of poor disease outcomes across all clinical outcomes assessed. Monocytes (percentage) and platelet counts also demonstrated strong prognostic value, though at baseline they were within normal ranges. Elevated LDH levels appeared to have prognostic value, but this finding was not robust across different sensitivity analyses. Our findings agree with the hypothesis that systemic hyperinflammation, tissue damage, and dysregulated immune responses are associated with poor disease outcomes in patients with severe COVID-19.^4,9,10^ Elevated D-dimer levels as a marker of coagulopathy showed weak and inconsistent prognostic signals in COVACTA. However, there were potential confounding factors for D-dimer assessment in our study, including higher levels among survivors in COVACTA compared with those reported in other studies^10^ (Appendix Table S4), a disproportionate balance of missing data by region (59.2% for Europe, 40.8% for North America), an association with the stratification factor of region that was not evident for any other biomarkers tested, and an unadjusted p=0.023 for mortality in prognostic modelling compared with an adjusted p=0.54 for region. Thus, the prognostic value of D-dimer levels in COVID-19 remains controversial.^12,21,22^

Ferritin was the only biomarker in this exploratory analysis that predicted the effect of tocilizumab treatment in COVACTA across multiple clinical outcomes. During infection and inflammation, ferritin is released from macrophages and hepatocytes to protect cells from oxidative damage and sequester iron from pathogens.^23,24^ Although IL-6 stimulates hepatocytes to release acute-phase proteins, including ferritin, as a systemic response, ferritin is also a marker of myeloid cell activation in the blood and inflamed tissue. Once released, it can further activate macrophages to produce cytokines such as IL-6 and IL-1β, driving a positive feedback loop of ferritin-inducing proinflammatory cytokine release.^23,24^ Altered myeloid cell activation and elevated ferritin levels correlate with increased severity of COVID-19 disease.^1,25^ Furthermore, elevated ferritin levels decreased significantly after tocilizumab administration in patients with severe COVID-19,^12^ suggesting a possible mechanism underlying the predictive value of ferritin for tocilizumab treatment effects. High ferritin levels are thought to be a biomarker and a driver of disease severity in other clinical disorders characterised by abnormal macrophage activation and elevated IL-6 activity, such as cytokine release syndrome^26^ and systemic juvenile idiopathic arthritis (sJIA).^27^ In both conditions, patients benefit from tocilizumab treatment, and ferritin levels decrease after treatment in those with sJIA.^27^ Although results are hypothesis-generating in this respect, the relationship between hyper-ferritinemia and elevated IL-6 levels in COVID-19 and other diseases provides a potential rationale for why ferritin was predictive for the effects of IL-6 pathway inhibition with tocilizumab in our study. Additional investigation of biomarkers downstream of IL-6 and ferritin signalling, together with single-cell analysis of pulmonary cells, might further elucidate how high ferritin levels predict benefit from the inhibition of IL-6 activity in patients with severe COVID-19.

IL-6 and CRP are inflammatory markers that respond to tocilizumab treatment.^28^ Although they demonstrated strong prognostic value as disease biomarkers in COVID-19, elevated levels of IL-6 or CRP at baseline were not predictive of the tocilizumab treatment effect in the overall COVACTA population. IL-6 only demonstrated a trend for predictive value among the subgroup of patients who required high-flow oxygen and non-invasive or invasive mechanical ventilation (ordinal scale categories 4 and 5) at baseline. This result might be counterintuitive given that tocilizumab inhibits the IL-6 receptor to block the IL-6 pathway; however, it is not entirely unexpected. Since approval of tocilizumab for the treatment of rheumatoid arthritis in 2009, no predictive biomarker—neither IL-6 nor CRP—has been validated for tocilizumab treatment effects in rheumatoid diseases.^28,29^ Similarly, despite the positive benefit/risk of tocilizumab treatment in other autoimmune indications, such as giant cell arteritis, the predictive value of baseline IL-6 or CRP has not been established.^29^ It remains unclear how IL-6 activity measured in blood is associated with tocilizumab efficacy endpoints in complex diseases; further investigation in clinical studies is warranted.

Because the COVACTA trial did not meet its primary endpoint of improved clinical status in patients with COVID-19–associated pneumonia or the key secondary endpoint of reduced patient mortality, identifying a subgroup of patients who might have derived clinical benefit in our study (patients with elevated ferritin levels at baseline) also reveals a subgroup potentially harmed (patients without highly elevated ferritin levels at baseline). This, combined with the preliminary nature of our results, means that extreme caution must be exercised and that additional study is required before a clinically useful numerical cut-off for ferritin levels can be defined. Another limitation of this study is that laboratory values were measured locally, which, notably, entails more variability than uniform test runs in a central laboratory. Local laboratory results for ferritin, however, were confirmed by retesting all samples at a central laboratory using an in vitro diagnostic method (Roche Cobas).

In conclusion, markers of hyperinflammation, macrophage activation, and dysregulated immune cells appear to be important prognostic biomarkers in COVID-19. Ferritin, as an acute-phase protein and a macrophage activation marker, might have potential as a predictive biomarker to help identify patients with severe COVID-19 who respond to tocilizumab. Results from ongoing studies of tocilizumab in COVID-19 are needed to validate these findings.

## Supporting information

APPENDIX for COVACTA Manuscript

## Data Availability

Qualified researchers may request access to individual patient level data through the clinical study data request platform (https://vivli.org/). Further details on Roche's criteria for eligible studies are available here (https://vivli.org/members/ourmembers/). For further details on Roche's Global Policy on the Sharing of Clinical Information and how to request access to related clinical study documents, see here (https://www.roche.com/research_and_development/who_we_are_how_we_work/clinical_trials/our_commitment_to_data_sharing.htm).    

https://vivli.org/members/ourmembers/

https://www.roche.com/research_and_development/who_we_are_how_we_work/clinical_trials/our_commitment_to_data_sharing.htm

## Contributors

Jennifer Tom contributed to data analysis and interpretation.

Min Bao contributed to study design and to data collection, analysis, and interpretation.

Larry Tsai contributed to study design and data interpretation.

Aditi Qamra and David Summers contributed to data analysis, figure creation, and data interpretation.

Montserrat Carrasco-Triguero contributed to biomarker analysis.

Jacqueline McBride contributed to study design and to data analysis and interpretation.

Carrie M Rosenberger contributed to the biomarker study design and data interpretation.

Celia J F Lin contributed to design of the MARIPOSA trial and to data analysis and interpretation.

William Stubbings contributed to the MARIPOSA trial clinical science team and to data analysis and interpretation of the MARIPOSA trial.

Kevin G Blyth contributed to enrolment of patients and to data collection and interpretation. Jordi Carratalà, Bruno François, and Paolo Bonfanti contributed to data collection.

Thomas Benfield and Cor H van der Leest contributed to enrolment of patients and data collection.

Derrick Haslem was a study site principal investigator.

Nidhi Rohatgi contributed to data collection and interpretation.

Charles Edouard Luyt contributed to enrolment of patients and data collection. Farrah Kehradmand contributed to data interpretation.

Fang Cai contributed to the literature search, generation of the biomarker strategy, planning and implementation of the biomarker analysis, and data collection, analysis, and interpretation.

All authors were involved in writing the manuscript or revising it critically for important intellectual content and approving the final version for publication, and all verify that they had access to and take responsibility for all the data in the manuscript.

## Acknowledgments

Medical writing support was provided by Sara Duggan, PhD, of ApotheCom, and was funded by F. Hoffmann-La Roche Ltd. The study was funded by F. Hoffmann-La Roche Ltd and, in part, by federal funds received from the US Department of Health and Human Services, Office of the Assistant Secretary for Preparedness and Response, Biomedical Advanced Research and Development Authority, under OT number HHSO100201800036C.

## References

1 Vabret N, Britton GJ, Gruber C, et al. Immunology of COVID-19: current state of the science. Immunity 2020; 52: 910–41.

2 Giamarellos-Bourboulis EJ, Netea MG, Rovina N, et al. Complex immune dysregulation in COVID-19 patients with severe respiratory failure. Cell Host Microbe 2020; 6: 992-1000.e3.

3 Mazzoni A, Salvati L, Maggi L, et al. Impaired immune cell cytotoxicity in severe COVID-19 is IL-6 dependent. J Clin Invest 2020; 130: 4694–703.

4 Zhu J, Pang J, Ji P, et al. Elevated interleukin-6 is associated with severity of COVID-19: a meta-analysis. J Med Virol 2020; 10.1002/jmv.26085. doi:10.1002/jmv.26085.

5 Kaye AG, Siegel R. The efficacy of IL-6 inhibitor tocilizumab in reducing severe COVID-19 mortality: a systematic review. medRxiv. 2020; 10.20150938.

6 Guaraldi G, Meschiari M, Cozzi-Lepri A, et al. Tocilizumab in patients with severe COVID-19: a retrospective cohort study. Lancet Rheumatol 2020; 2: e474–e84.

7 Rosas IO, Bräu N, Waters M, et al. Tocilizumab in hospitalized patients with COVID-19 pneumonia. medRxiv 2020; 10.1101/2020.08.27.20183442.

8 National Insitutes of Health. COVID-19 treatment guidelines: coronavirus disease 2019 (COVID-19) treatment guidelines. https://www.covid19treatmentguidelines.nih.gov/. Accessed November 16, 2020.

9 Herold T, Jurinovic V, Arnreich C, et al. Elevated levels of IL-6 and CRP predict the need for mechanical ventilation in COVID-19. J Allergy Clin Immunol 2020; 146: 128-36.e4.

10 Zhou F, Yu T, Du R, et al. Clinical course and risk factors for mortality of adult inpatients with COVID-19 in Wuhan, China: a retrospective cohort study. Lancet 2020; 395: 1054–62.

11 Lohse A, Klopfenstein T, Balblanc JC, et al. Predictive factors of mortality in patients treated with tocilizumab for acute respiratory distress syndrome related to coronavirus disease 2019 (COVID-19). Microbes Infect 2020; 22: 500–3.

12 Conrozier T, Lohse A, Balblanc JC, et al. Biomarker variation in patients successfully treated with tocilizumab for severe coronavirus disease 2019 (COVID-19): results of a multidisciplinary collaboration. Clin Exp Rheumatol 2020; 38: 742–7.

13 Luo P, Liu Y, Qiu L, Liu X, Liu D, Li J. Tocilizumab treatment in COVID-19: a single center experience. J Med Virol 2020; 92: 814–8.

14 Lok LSC, Farahi N, Juss JK, et al. Effects of tocilizumab on neutrophil function and kinetics. Eur J Clin Invest 2017; 47: 736–45.

15 Gibiansky L, Frey N. Linking interleukin-6 receptor blockade with tocilizumab and its hematological effects using a modeling approach. J Pharmacokinet Pharmacodyn 2012; 39: 5–16.

16 Fonseca JE, Santos MJ, Canhao H, Choy E. Interleukin-6 as a key player in systemic inflammation and joint destruction. Autoimmun Rev 2009; 8: 538–42.

17 Li F, Morgan KL, Zaslavsky AM. Balancing covariates via propensity score weighting. J Am Stat Assoc 2018; 113: 390–400.

18 Stuart EA, Lee BK, Leacy FP. Prognostic score-based balance measures can be a useful diagnostic for propensity score methods in comparative effectiveness research. J Clin Epidemiol 2013; 66(suppl): S84-90.e1.

19 Austin PC, Stuart EA. Moving towards best practice when using inverse probability of treatment weighting (IPTW) using the propensity score to estimate causal treatment effects in observational studies. Stat Med 2015; 34: 3661–79.

20 Henry BM, de Oliveira MHS, Benoit S, Plebani M, Lippi G. Hematologic, biochemical and immune biomarker abnormalities associated with severe illness and mortality in coronavirus disease 2019 (COVID-19): a meta-analysis. Clin Chem Lab Med 2020; 58: 1021–8.

21 Ye W, Chen G, Li X, et al. Dynamic changes of D-dimer and neutrophil-lymphocyte count ratio as prognostic biomarkers in COVID-19. Respir Res 2020; 21: 169.

22 Leisman DE, Ronner L, Pinotti R, et al. Cytokine elevation in severe and critical COVID-19: a rapid systematic review, meta-analysis, and comparison with other inflammatory syndromes. Lancet Respir Med 2020; 8: 1233–44.

23 Kappert K, Jahić A, Tauber R. Assessment of serum ferritin as a biomarker in COVID-19: bystander or participant? Insights by comparison with other infectious and non-infectious diseases. Biomarkers 2020: 1–10. 10.1080/1354750X.2020.1797880.

24 Ruscitti P, Di Benedetto P, Berardicurti O, et al. Pro-inflammatory properties of H-ferritin on human macrophages, ex vivo and in vitro observations. Sci Rep 2020; 10: 12232.

25 Schulte-Schrepping J, Reusch N, Paclik D, et al. Severe COVID-19 is marked by a dysregulated myeloid cell compartment. Cell 2020; 182: 1419-40.e23.

26 Teachey DT, Lacey SF, Shaw PA, et al. Identification of predictive biomarkers for cytokine release syndrome after chimeric antigen receptor T-cell therapy for acute lymphoblastic leukemia. Cancer Discov 2016; 6: 664–79.

27 Shimizu M, Mizuta M, Okamoto N, et al. Tocilizumab modifies clinical and laboratory features of macrophage activation syndrome complicating systemic juvenile idiopathic arthritis. Pediatr Rheumatol Online J 2020; 18: 2.

28 Wang J, Platt A, Upmanyu R, et al. IL-6 pathway-driven investigation of response to IL-6 receptor inhibition in rheumatoid arthritis. BMJ Open 2013; 3: e003199.

29 Choy EH, De Benedetti F, Takeuchi T, Hashizume M, John MR, Kishimoto T. Translating IL-6 biology into effective treatments. Nat Rev Rheumatol 2020; 16: 335–45.

30 Tang N, Li D, Wang X, Sun Z. Abnormal coagulation parameters are associated with poor prognosis in patients with novel coronavirus pneumonia. J Thromb Haemost 2020; 18: 844–7.

