## APPENDIX for COVACTA Manuscript for "Prognostic and predictive biomarkers in patients with COVID-19 treated with tocilizumab in a randomised controlled trial"

### SUPPLEMENTARY APPENDIX

***Appendix 1.*** **Institutional review boards and ethics committees**

| | | |
|---|---|---|
| COMITATO ETICO DELL'IRCCS SAN MATTEO DI PAVIA | COMITATO ETICO DELL'IRCCS SAN MATTEO DI PAVIA, IRCCS Policlinico San Matteo, Viale Golgi ,19, 27100, Pavia, Lombardia, ITALY | ITALY |
| Advarra | Advarra, 6940 Columbia Gateway Drive, COLUMBIA, MD, 21046, UNITED STATES | UNITED STATES |
| CPP Sud Ouest Et Outre Mer I | CPP Sud Ouest Et Outre Mer I, 10 chemin du raisin, ARS Midi-Pyrénées, Bureau 1028, 31050, Toulouse cedex 9, FRANCE | FRANCE |
| CPP Sud Ouest Et Outre Mer I | CPP Sud Ouest Et Outre Mer I, 10 chemin du raisin, ARS Midi-Pyrénées, Bureau 1028, 31050, Toulouse cedex 9, FRANCE | FRANCE |
| CEIC Área 5 - Hospital Universitario La Paz | CEIC Área 5 - Hospital Universitario La Paz, Paseo de la Castellana, 261, 28036, Madrid, MADRID, SPAIN | SPAIN |
| CEIC Área 5 - Hospital Universitario La Paz | CEIC Área 5 - Hospital Universitario La Paz, Paseo de la Castellana, 261, 28036, Madrid, MADRID, SPAIN | SPAIN |
| CEIC Área 5 - Hospital Universitario La Paz | CEIC Área 5 - Hospital Universitario La Paz, Paseo de la Castellana, 261, 28036, Madrid, MADRID, SPAIN | SPAIN |
| CEIC Área 5 - Hospital Universitario La Paz | CEIC Área 5 - Hospital Universitario La Paz, Paseo de la Castellana, 261, 28036, Madrid, MADRID, SPAIN | SPAIN |
| Comité d'éthique de la recherche | Comité d'éthique de la recherche, CHUM, Pavillion R, 900 Rue St-Denis, 3rd Floor, H2X 0A9, Montreal, Quebec, CANADA | CANADA |
| Advarra | Advarra, 6940 Columbia Gateway Drive, COLUMBIA, MD, 21046, UNITED STATES | UNITED STATES |
| Advarra | Advarra, 6940 Columbia Gateway Drive, COLUMBIA, MD, 21046, UNITED STATES | |
| Advarra | Advarra, 6940 Columbia Gateway Drive, COLUMBIA, MD, 21046, UNITED STATES | UNITED STATES |
| Stichting Beoordeling Ethiek Biomedisch Onderzoek (BEBO) | Stichting Beoordeling Ethiek Biomedisch Onderzoek (BEBO), Dr. Nassaulaan 10, 9401 HK, Assen, NETHERLANDS | NETHERLANDS |
| Stichting Beoordeling Ethiek Biomedisch Onderzoek (BEBO) | Stichting Beoordeling Ethiek Biomedisch Onderzoek (BEBO), Dr. Nassaulaan 10, 9401 HK, Assen, NETHERLANDS | NETHERLANDS |
| West Midlands - Coventry & Warwickshire Research Ethics Committee | West Midlands - Coventry & Warwickshire Research Ethics Committee, The Old Chapel, Royal Standard Place, Nottingham, NG1 6FS, UNITED KINGDOM | UNITED KINGDOM |
| Ethikkommission der Medizinischen Fakultät Universität zu Köln | Ethikkommission der Medizinischen Fakultät Universität zu Köln, Kerpener Str.62, Gebäude 5, 50937, Köln, GERMANY | GERMANY |
| Ethikkommission der Medizinischen Fakultät Universität zu Köln | Ethikkommission der Medizinischen Fakultät Universität zu Köln, Kerpener Str.62, Gebäude 5, 50937, Köln, GERMANY | GERMANY |

| | | |
|---|---|---|
| Ethikkommission der Medizinischen Fakultät Universität zu Köln | Ethikkommission der Medizinischen Fakultät Universität zu Köln, Kerpener Str.62, Gebäude 5, 50937, Köln, GERMANY | GERMANY |
| Advarra | Advarra, 6940 Columbia Gateway Drive, COLUMBIA, MD, 21046, UNITED STATES | UNITED STATES |
| University Health Network Research Ethics Board | University Health Network Research Ethics Board, 700 Bay Street, 17th Floor, Suite 1700, M5G 1Z6, Toronto, Ontario, CANADA | CANADA |
| CPP Sud Ouest Et Outre Mer I | CPP Sud Ouest Et Outre Mer I, 10 chemin du raisin, ARS Midi-Pyrénées, Bureau 1028, 31050, Toulouse cedex 9, FRANCE | FRANCE |
| CPP Sud Ouest Et Outre Mer I | CPP Sud Ouest Et Outre Mer I, 10 chemin du raisin, ARS Midi-Pyrénées, Bureau 1028, 31050, Toulouse cedex 9, FRANCE | FRANCE |
| CPP Sud Ouest Et Outre Mer I | CPP Sud Ouest Et Outre Mer I, 10 chemin du raisin, ARS Midi-Pyrénées, Bureau 1028, 31050, Toulouse cedex 9, FRANCE | FRANCE |
| CPP Sud Ouest Et Outre Mer I | CPP Sud Ouest Et Outre Mer I, 10 chemin du raisin, ARS Midi-Pyrénées, Bureau 1028, 31050, Toulouse cedex 9, FRANCE | FRANCE |
| Videnskabsetiske Komité Region Midt | Videnskabsetiske Komité Region Midt; Sundhedssekr., Skottenborg 26, Postboks 21, 8800, Viborg, DENMARK | DENMARK |
| Videnskabsetiske Komité Region Midt | Videnskabsetiske Komité Region Midt; Sundhedssekr., Skottenborg 26, Postboks 21, 8800, Viborg, DENMARK | DENMARK |
| Videnskabsetiske Komité Region Midt | Videnskabsetiske Komité Region Midt; Sundhedssekr., Skottenborg 26, Postboks 21, 8800, Viborg, DENMARK | DENMARK |
| Videnskabsetiske Komité Region Midt | Videnskabsetiske Komité Region Midt; Sundhedssekr., Skottenborg 26, Postboks 21, 8800, Viborg, DENMARK | DENMARK |
| CPP Sud Ouest Et Outre Mer | CPP Sud Ouest Et Outre Mer I, 10 chemin du raisin, ARS Midi-Pyrénées, Bureau 1028, 31050, Toulouse cedex 9, FRANCE | FRANCE |
| COMITATO ETICO BRIANZA | COMITATO ETICO BRIANZA, Via Pergolesi, 33, 20900, Monza, Lombardia, ITALY | ITALY |
| Hamilton Integrated Research Ethics Board | Hamilton Integrated Research Ethics Board, 293 Wellington St. North, Suite 102, L8L 8E7, Hamilton, Ontario, CANADA | CANADA |
| Hamilton Integrated Research Ethics Board | Hamilton Integrated Research Ethics Board, 293 Wellington St. North, Suite 102, L8L 8E7, Hamilton, Ontario, CANADA | CANADA |
| Comitato Etico della Seconda Università di Napoli; Az. Osp. Univ. S.U.N. - A.O.R.N. "Ospedali | Comitato Etico della Seconda Università di Napoli; Az. Osp. Univ. S.U.N. - A.O.R.N. "Ospedali, Via Costantinopoli, 104, 80138,Napoli, ITALY | ITALY |
| Comitato Etico Ist. Naz. Mal. Inf. L. Spallanzani | Comitato Etico Ist. Naz. Mal. Inf. L. Spallanzani, Via Portuense, 292, 149, Roma, ITALY | ITALY |
| COMITATO ETICO DELLA PROVINCIA DI BERGAMO | COMITATO ETICO DELLA PROVINCIA DI BERGAMO, Piazza OMS-Organizzazione Mondiale della Sanità, 1, 24127, Bergamo,ITALY | ITALY |

| | | |
|---|---|---|
| COMITATO ETICO MILANO AREA 1; c/o ASST FBF Sacco - P.O. L. Sacco | COMITATO ETICO MILANO AREA 1; c/o ASST FBF Sacco - P.O. L. Sacco, Via G.B. Grassi, 74, 20157, MILANO, ITALY | ITALY |
| Hamilton Integrated Research Ethics Board | Hamilton Integrated Research Ethics Board, 293 Wellington St. North, Suite 102, L8L 8E7, Hamilton, CANADA | CANADA |
| Stichting Beoordeling Ethiek Biomedisch Onderzoek (BEBO) | Stichting Beoordeling Ethiek Biomedisch Onderzoek (BEBO), Dr. Nassaulaan 10, 9401 HK, Assen, NETHERLANDS | NETHERLANDS |
| Stichting Beoordeling Ethiek Biomedisch Onderzoek (BEBO) | Stichting Beoordeling Ethiek Biomedisch Onderzoek (BEBO), Dr. Nassaulaan 10, 9401 HK, Assen, NETHERLANDS | NETHERLANDS |
| CEIC Área 5 - Hospital Universitario La Paz | CEIC Área 5 - Hospital Universitario La Paz, Paseo de la Castellana, 261, 28036, Madrid, MADRID, SPAIN | SPAIN |
| CEIC Área 5 - Hospital Universitario La Paz | CEIC Área 5 - Hospital Universitario La Paz, Paseo de la Castellana, 261, 28036, Madrid, MADRID, SPAIN | SPAIN |
| CEIC Área 5 - Hospital Universitario La Paz | CEIC Área 5 - Hospital Universitario La Paz, Paseo de la Castellana, 261, 28036, Madrid, MADRID, SPAIN | SPAIN |
| West Midlands - Coventry & Warwickshire Research Ethics Committee | West Midlands - Coventry & Warwickshire Research Ethics Committee, The Old Chapel, Royal Standard Place, Nottingham, NG1 6FS, UNITED KINGDOM | UNITED KINGDOM |
| West Midlands - Coventry & Warwickshire Research Ethics Committee | West Midlands - Coventry & Warwickshire Research Ethics Committee, The Old Chapel, Royal Standard Place, Nottingham, NG1 6FS, UNITED KINGDOM | UNITED KINGDOM |
| West Midlands - Coventry & Warwickshire Research Ethics Committee | West Midlands - Coventry & Warwickshire Research Ethics Committee, The Old Chapel, Royal Standard Place, Nottingham, NG1 6FS, UNITED KINGDOM | UNITED KINGDOM |
| West Midlands - Coventry & Warwickshire Research Ethics Committee | West Midlands - Coventry & Warwickshire Research Ethics Committee, The Old Chapel, Royal Standard Place, Nottingham, NG1 6FS, UNITED KINGDOM | UNITED KINGDOM |
| West Midlands - Coventry & Warwickshire Research Ethics Committee | West Midlands - Coventry & Warwickshire Research Ethics Committee, The Old Chapel, Royal Standard Place, Nottingham, NG1 6FS, UNITED KINGDOM | UNITED KINGDOM |
| West Midlands - Coventry & Warwickshire Research Ethics Committee | West Midlands - Coventry & Warwickshire Research Ethics Committee, The Old Chapel, Royal Standard Place, Nottingham, NG1 6FS, UNITED KINGDOM | UNITED KINGDOM |
| West Midlands - Coventry & Warwickshire Research Ethics Committee | West Midlands - Coventry & Warwickshire Research Ethics Committee, The Old Chapel, Royal Standard Place, Nottingham, NG1 6FS, UNITED KINGDOM | UNITED KINGDOM |
| Advarra | Advarra, 6940 Columbia Gateway Drive, COLUMBIA, MD, 21046, UNITED STATES | UNITED STATES |
| Advarra | Advarra, 6940 Columbia Gateway Drive, COLUMBIA, MD, 21046, UNITED STATES | UNITED STATES |

| | | |
|---|---|---|
| Advarra | Advarra, 6940 Columbia Gateway Drive, COLUMBIA, MD, 21046, UNITED STATES | UNITED STATES |
| Advarra | Advarra, 6940 Columbia Gateway Drive, COLUMBIA, MD, 21046, UNITED STATES | UNITED STATES |
| Advarra | Advarra, 6940 Columbia Gateway Drive, COLUMBIA, MD, 21046, UNITED STATES | UNITED STATES |
| Advarra | Advarra, 6940 Columbia Gateway Drive, COLUMBIA, MD, 21046, UNITED STATES | UNITED STATES |
| Advarra | Advarra, 6940 Columbia Gateway Drive, COLUMBIA, MD, 21046, UNITED STATES | UNITED STATES |
| Sharp HealthCare | Sharp HealthCare, IRB, 7930 Frost Street, Suite 300, SAN DIEGO, CA, 92123, UNITED STATES | UNITED STATES |
| Rush University Medical Center | Rush University Medical Center; Rush University Medical Center Institutional Review Board, 1653 West Congress Parkway, Jelke Building Room 1591, Chicago, IL, 60612, UNITED STATES | UNITED STATES |
| Cleveland Clinic | Cleveland Clinic; Institutional Review Board, 10681 Carnegie Ave, OS-1, Cleveland, OH, 44195, UNITED STATES | UNITED STATES |
| Advarra | Advarra, 6940 Columbia Gateway Drive, COLUMBIA, MD, 21046, UNITED STATES | UNITED STATES |
| Advarra | Advarra, 6940 Columbia Gateway Drive, COLUMBIA, MD, 21046, UNITED STATES | UNITED STATES |
| Providence St. Joseph Health IRB | Providence St. Joseph Health IRB, 1801 Lind Ave SW, RENTON, WA, 98057, UNITED STATES | UNITED STATES |
| Advarra | Advarra, 6940 Columbia Gateway Drive, COLUMBIA, MD, 21046, UNITED STATES | UNITED STATES |
| Duke University | Duke University; DUHS Institutional Review Board, Hock Plaza, 2424 Erwin Rd, Durham, NC, UNITED STATES | UNITED STATES |
| Bronx VAMC IRB | Bronx VAMC IRB, 130 West Kingsbridge Road, 2Fo1 Research & Development, Bronx, NY, 10468, UNITED STATES | UNITED STATES |
| Advarra | Advarra, 6940 Columbia Gateway Drive, COLUMBIA, MD, 21046, UNITED STATES | UNITED STATES |
| Univ of Chicago | Univ of Chicago; Institutional Review Board, 5841 S. Maryland Ave., MC7132, I-625, CHICAGO, IL, 60637, UNITED STATES | UNITED STATES |

*Appendix Table S1:* **Baseline characteristics of patients in MARIPOSA (severe disease subset) and COVACTA**

| | **COVACTA N=437** | **MARIPOSA N=77** |
|---|---|---|
| Mechanical ventilation, n (%) | | |
| No | 270 (61·8) | 65 (84·4) |
| Yes | 167 (38·2) | 12 (15·6) |
| Ordinal scale category, n (%) | | |
| 2 | 15 (3·4) | 2 (2·6) |
| 3 | 122 (27·9) | 22 (28·6) |
| 4 | 133 (30·4) | 41 (53·2) |
| 5 | 60 (13·7) | 9 (11·7) |
| 6 | 107 (24·5) | 3 ( 3·9) |
| Sex, n (%) | | |
| Female | 136 (30·2) | 29 (37·7) |
| Male | 315 (69·8) | 48 (62·3) |
| Age | | |
| Mean (SD) | 61·0 (14·3) | 59·1 (14·4) |
| Median | 63 | 60 |
| Q1–Q3 | 53–71 | 50–70 |
| Min–Max | 22–96 | 27–91 |
| Antiviral use, n (%) | | |
| No | 336 (74·5) | 41 (53·2) |
| Yes | 115 (25·5) | 36 (46·8) |
| Steroid use, n (%) | | |
| No | 353 (78·3) | 57 (74·0) |
| Yes | 98 (21·7) | 20 (26·0) |

One patient who had an ordinal scale score of 7 at baseline (death) was excluded from the analysis.

Q=quarter.

*Appendix Table S2:* **Combinations of predictive biomarkers**

| **Biomarker Combination** | **Estimate** | **p-value** | **2·5%** | **97·5%** |
|---|---|---|---|---|
| **Combined ferritin, IL-6, and CRP (n=257)** | | | | |
| Ferritin | 0·20 | 0·04 | 0·04 | 0·87 |
| IL-6 | 0·89 | 0·88 | 0·18 | 3·97 |
| CRP | 0·48 | 0·42 | 0·06 | 2·51 |
| Ferritin and IL-6 | 0·62 | 0·52 | 0·14 | 2·69 |
| Ferritin and CRP | 0·41 | 0·23 | 0·09 | 1·73 |
| IL-6 and CRP | 0·79 | 0·75 | 0·18 | 3·32 |
| Ferritin or IL-6 | 0·48 | 0·16 | 0·16 | 1·30 |
| Ferritin or CRP | 0·40 | 0·09 | 0·12 | 1·08 |
| IL-6 or CRP | 0·82 | 0·68 | 0·29 | 2·09 |
| **Combined ferritin, LDH, and D-dimer (n=165)** | | | | |
| Ferritin | 0·19 | 0·05 | 0·03 | 0·99 |
| D-dimer | 1·16 | 0·86 | 0·22 | 6·09 |
| LDH | 1·68 | 0·56 | 0·29 | 9·61 |
| Ferritin and D-dimer | 0·54 | 0·49 | 0·09 | 3·13 |
| Ferritin and LDH | 0·32 | 0·20 | 0·05 | 1·83 |
| D-dimer and LDH | 1·97 | 0·44 | 0·36 | 11·46 |
| Ferritin or D-dimer | 0·51 | 0·25 | 0·16 | 1·56 |
| Ferritin or LDH | 0·47 | 0·22 | 0·14 | 1·51 |
| D-dimer or LDH | 1·30 | 0·66 | 0·40 | 4·07 |
| LDH and D-dimer and ferritin | 0·50 | 0·50 | 0·06 | 3·93 |

A median cut-off value was used for each biomarker.

CRP=C-reactive protein; IL-6=interleukin-6; LDH=lactate dehydrogenase.

***Appendix Table S3:*** **Ferritin levels according to baseline ordinal scale**

| **Baseline ordinal scale category** | **Mean** | **Median** | **25%** | **75%** | **Standard Deviation** | **n** |
|---|---|---|---|---|---|---|
| 2 | 1135·88 | 640·17 | 201·78 | 2071·73 | 1036·74 | 12 |
| 3 | 2126·42 | 1677·39 | 870·15 | 2798·08 | 1634·71 | 104 |
| 4 | 3978·09 | 2422·27 | 1256·07 | 4458·50 | 7849·37 | 103 |
| 5 | 4315·15 | 3148·05 | 1659·09 | 5207·03 | 3860·48 | 54 |
| 6 | 3619·02 | 2464·96 | 1297·69 | 4392·89 | 4132·11 | 91 |

Ferritin levels are shown as pmol/L.

*Appendix Table S4:* **D-dimer levels among survivors and non-survivors in COVACTA and published studies**

| | Non-survivors | | Survivors | |
|---|---|---|---|---|
| D-dimer, µg/mL* | **N** | **Mean (IQR) [median]** | **N** | **Mean (IQR) [median]** |
| COVACTA | 47 | 7·16 (1·02–8·37) [2·68] | 150 | 3·58 (0·63–2·32) [1·13] |
| Zhou F et al 2020[10] | 54 | 5·2 (1·5–21·1) [NR] | 137 | 0·6 (0·3–1·0) [NR] |
| Tang N et al 2020[30] | 21 | 2·12 (0·77–5·27) [NR] | 162 | 0·61 (0·35–1·29) [NR] |

*Reported as fibrinogen equivalent units for COVACTA, though published studies do not state whether D-dimer was reported as fibrinogen equivalent units or D-dimer units.

IQR=interquartile range; NR=not reported.

*Appendix Figure S1:* **Baseline biomarker levels**

(A) Overall and (B) by treatment arm. (C) Correlation between baseline biomarkers.

**A**

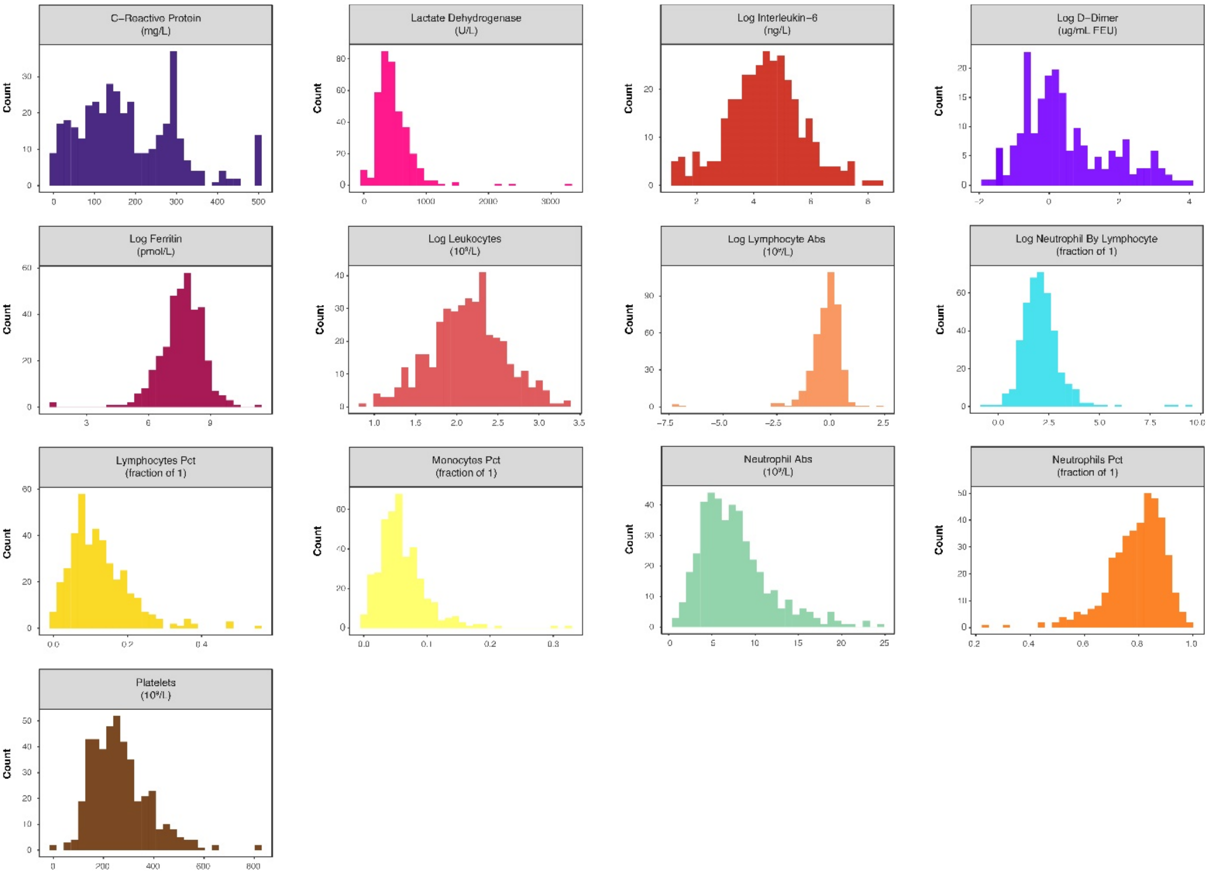

**B**

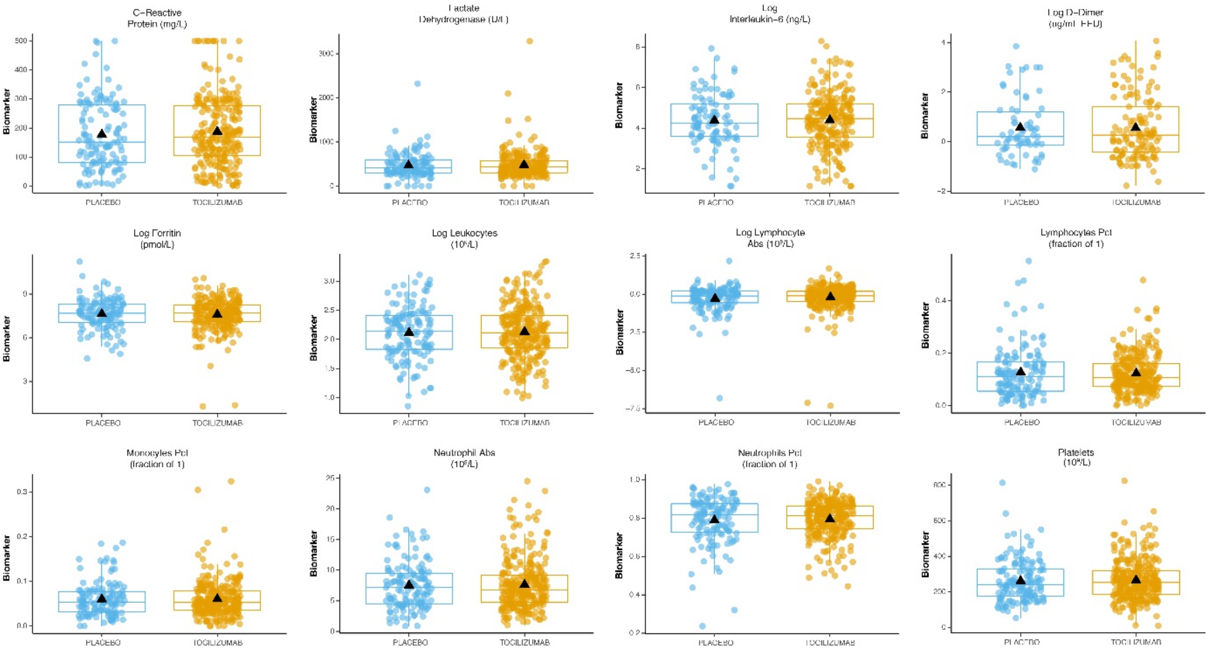
C−Reactive Protein (mg/L)
Lactate Dehydrogenase (U/L)
Log Interleukin−6 (ng/L)
Log D−Dimer (ug/mL FEU)
Log Ferritin (pmol/L)
Log Leukocytes (10⁹/L)
Log Lymphocyte Abs (10⁹/L)
Lymphocytes Pct (fraction of 1)
Monocytes Pct (fraction of 1)
Neutrophil Abs (10⁹/L)
Neutrophils Pct (fraction of 1)
Platelets (10⁹/L)
Biomarker
PLACEBO
TOCILIZUMAB

**C**

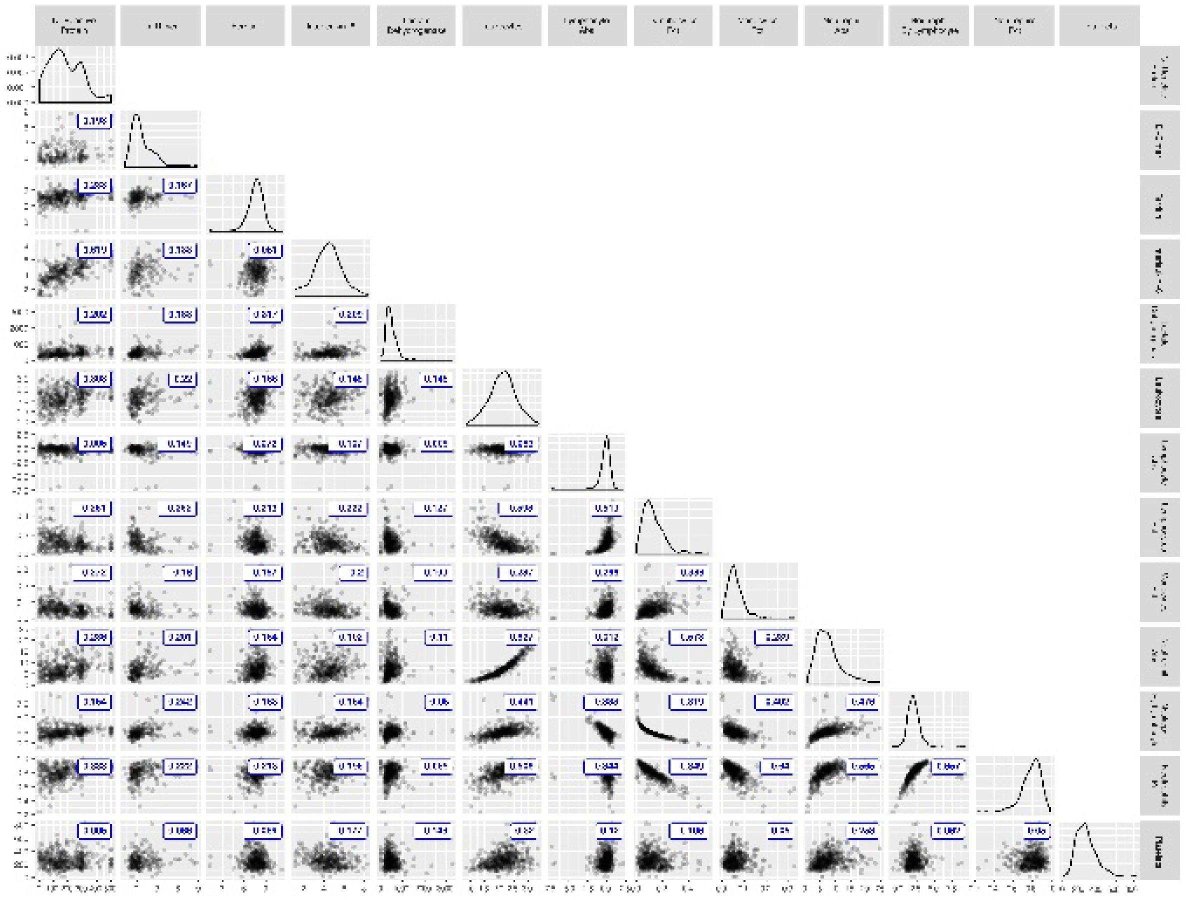

Ferritin, neutrophil-to-lymphocyte ratio, IL-6, D-dimer, absolute lymphocyte, and absolute leukocyte values are log transformed.

IL-6=interleukin-6.

*Appendix Figure S2:* **Prognostic forest plots**

(A) Ordinal scale and (B) time to hospital discharge.

**A**

**Ordinal Scale Day 28; Prognostic; Scaled & Adjusted**

| Biomarker | Model | Odds Ratio | Lower | Upper | N | P–Value |
|---|---|---|---|---|---|---|
| CRP | All | 1.54 | 1.27 | 1.86 | 382 | 0 |
| | All, Adjusted | 1.44 | 1.16 | 1.78 | 382 | 0 |
| | PBO | 1.82 | 1.3 | 2.58 | 126 | 0 |
| | PBO, Adjusted | 1.77 | 1.21 | 2.62 | 126 | 0 |
| log(D–Dimer) | All | 1.63 | 1.22 | 2.19 | 197 | 0 |
| | All, Adjusted | 1.28 | 0.93 | 1.77 | 197 | 0.13 |
| | PBO | 1.71 | 1.04 | 2.9 | 66 | 0.04 |
| | PBO, Adjusted | 1.11 | 0.58 | 2.08 | 66 | 0.75 |
| log(Ferritin) | All | 1.27 | 1.04 | 1.57 | 364 | 0.02 |
| | All, Adjusted | 1.16 | 0.94 | 1.47 | 364 | 0.19 |
| | PBO | 1.91 | 1.33 | 2.81 | 124 | 0 |
| | PBO, Adjusted | 1.68 | 1.12 | 2.55 | 124 | 0.01 |
| log(IL6) | All | 1.94 | 1.56 | 2.45 | 332 | 0 |
| | All, Adjusted | 1.83 | 1.44 | 2.35 | 332 | 0 |
| | PBO | 1.86 | 1.26 | 2.83 | 99 | 0 |
| | PBO, Adjusted | 2.06 | 1.31 | 3.37 | 99 | 0 |
| LDH | All | 1.41 | 1.16 | 1.75 | 364 | 0 |
| | All, Adjusted | 1.65 | 1.34 | 2.09 | 364 | 0 |
| | PBO | 1.19 | 0.87 | 1.66 | 121 | 0.28 |
| | PBO, Adjusted | 1.3 | 0.9 | 1.93 | 121 | 0.17 |
| log(Lymphocytes) | All | 0.68 | 0.54 | 0.83 | 426 | 0 |
| | All, Adjusted | 0.81 | 0.66 | 0.98 | 426 | 0.03 |
| | PBO | 0.53 | 0.36 | 0.75 | 139 | 0 |
| | PBO, Adjusted | 0.71 | 0.47 | 0.97 | 139 | 0.05 |
| % Lymphocytes | All | 0.39 | 0.29 | 0.51 | 401 | 0 |
| | All, Adjusted | 0.53 | 0.4 | 0.7 | 401 | 0 |
| | PBO | 0.33 | 0.21 | 0.51 | 133 | 0 |
| | PBO, Adjusted | 0.5 | 0.3 | 0.78 | 133 | 0 |
| % Monocytes | All | 0.67 | 0.53 | 0.83 | 400 | 0 |
| | All, Adjusted | 0.65 | 0.51 | 0.82 | 400 | 0 |
| | PBO | 0.44 | 0.28 | 0.66 | 131 | 0 |
| | PBO, Adjusted | 0.44 | 0.27 | 0.69 | 131 | 0 |
| Neutrophils | All | 1.77 | 1.47 | 2.14 | 430 | 0 |
| | All, Adjusted | 1.45 | 1.18 | 1.79 | 430 | 0 |
| | PBO | 1.96 | 1.42 | 2.77 | 139 | 0 |
| | PBO, Adjusted | 1.56 | 1.07 | 2.3 | 139 | 0.02 |
| % Neutrophils | All | 1.98 | 1.57 | 2.51 | 402 | 0 |
| | All, Adjusted | 1.71 | 1.35 | 2.2 | 402 | 0 |
| | PBO | 2.51 | 1.72 | 3.81 | 134 | 0 |
| | PBO, Adjusted | 2.15 | 1.44 | 3.37 | 134 | 0 |
| log(%Neut/%Lym) | All | 2.01 | 1.6 | 2.57 | 397 | 0 |
| | All, Adjusted | 1.49 | 1.2 | 1.9 | 397 | 0 |
| | PBO | 2.4 | 1.68 | 3.57 | 132 | 0 |
| | PBO, Adjusted | 1.69 | 1.17 | 2.6 | 132 | 0.01 |
| Platelets | All | 0.73 | 0.6 | 0.88 | 437 | 0 |
| | All, Adjusted | 0.69 | 0.55 | 0.84 | 437 | 0 |
| | PBO | 0.65 | 0.46 | 0.9 | 142 | 0.01 |
| | PBO, Adjusted | 0.55 | 0.38 | 0.78 | 142 | 0 |
| log(WBC) | All | 1.82 | 1.5 | 2.23 | 420 | 0 |
| | All, Adjusted | 1.52 | 1.22 | 1.9 | 420 | 0 |
| | PBO | 1.94 | 1.4 | 2.73 | 140 | 0 |
| | PBO, Adjusted | 1.54 | 1.04 | 2.32 | 140 | 0.03 |

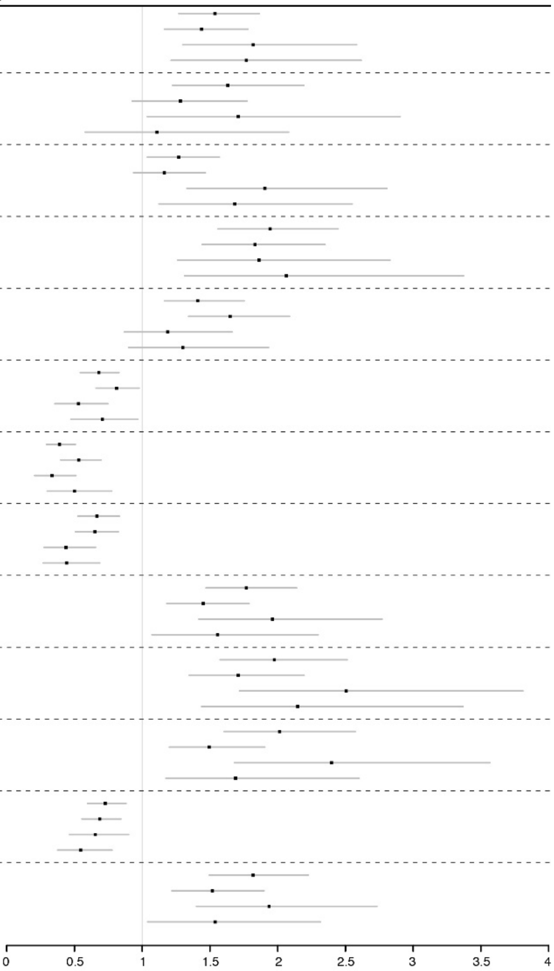

**B**

#### Time to Hospital Discharge (Fine–Gray); Prognostic; Scaled & Adjusted

| Biomarker | Model | Hazard Ratio | Lower | Upper | N | P–Value |
|---|---|---|---|---|---|---|
| CRP | All | 0.68 | 0.6 | 0.78 | 382 | 0 |
| | All, Adjusted | 0.73 | 0.63 | 0.85 | 382 | 0 |
| | PBO | 0.58 | 0.44 | 0.75 | 126 | 0 |
| | PBO, Adjusted | 0.61 | 0.47 | 0.8 | 126 | 0 |
| log(D–Dimer) | All | 0.67 | 0.54 | 0.84 | 197 | 0 |
| | All, Adjusted | 0.83 | 0.68 | 1.02 | 197 | 0.08 |
| | PBO | 0.68 | 0.44 | 1.06 | 66 | 0.09 |
| | PBO, Adjusted | 0.84 | 0.57 | 1.25 | 66 | 0.4 |
| log(Ferritin) | All | 0.83 | 0.72 | 0.96 | 364 | 0.02 |
| | All, Adjusted | 0.84 | 0.73 | 0.97 | 364 | 0.02 |
| | PBO | 0.65 | 0.53 | 0.81 | 124 | 0 |
| | PBO, Adjusted | 0.74 | 0.58 | 0.93 | 124 | 0.01 |
| log(IL6) | All | 0.64 | 0.56 | 0.74 | 332 | 0 |
| | All, Adjusted | 0.56 | 0.47 | 0.68 | 332 | 0 |
| | PBO | 0.6 | 0.43 | 0.83 | 99 | 0 |
| | PBO, Adjusted | 0.47 | 0.3 | 0.73 | 99 | 0 |
| LDH | All | 0.71 | 0.59 | 0.85 | 364 | 0 |
| | All, Adjusted | 0.6 | 0.48 | 0.75 | 364 | 0 |
| | PBO | 0.85 | 0.65 | 1.12 | 121 | 0.25 |
| | PBO, Adjusted | 0.81 | 0.58 | 1.13 | 121 | 0.21 |
| log(Lymphocytes) | All | 1.44 | 1.16 | 1.8 | 426 | 0 |
| | All, Adjusted | 1.4 | 1.08 | 1.83 | 426 | 0.01 |
| | PBO | 2.14 | 1.66 | 2.76 | 139 | 0 |
| | PBO, Adjusted | 1.73 | 1.32 | 2.28 | 139 | 0 |
| % Lymphocytes | All | 1.63 | 1.45 | 1.82 | 401 | 0 |
| | All, Adjusted | 1.46 | 1.32 | 1.63 | 401 | 0 |
| | PBO | 1.69 | 1.4 | 2.05 | 133 | 0 |
| | PBO, Adjusted | 1.49 | 1.26 | 1.77 | 133 | 0 |
| % Monocytes | All | 1.19 | 1.04 | 1.36 | 400 | 0.01 |
| | All, Adjusted | 1.19 | 1 | 1.41 | 400 | 0.05 |
| | PBO | 1.64 | 1.31 | 2.05 | 131 | 0 |
| | PBO, Adjusted | 1.69 | 1.36 | 2.1 | 131 | 0 |
| Neutrophils | All | 0.59 | 0.49 | 0.69 | 430 | 0 |
| | All, Adjusted | 0.73 | 0.61 | 0.87 | 430 | 0 |
| | PBO | 0.42 | 0.3 | 0.59 | 139 | 0 |
| | PBO, Adjusted | 0.54 | 0.37 | 0.77 | 139 | 0 |
| % Neutrophils | All | 0.66 | 0.58 | 0.74 | 402 | 0 |
| | All, Adjusted | 0.68 | 0.6 | 0.76 | 402 | 0 |
| | PBO | 0.6 | 0.49 | 0.74 | 134 | 0 |
| | PBO, Adjusted | 0.64 | 0.53 | 0.76 | 134 | 0 |
| log(%Neut/%Lym) | All | 0.5 | 0.41 | 0.61 | 397 | 0 |
| | All, Adjusted | 0.6 | 0.48 | 0.75 | 397 | 0 |
| | PBO | 0.41 | 0.31 | 0.55 | 132 | 0 |
| | PBO, Adjusted | 0.51 | 0.39 | 0.67 | 132 | 0 |
| Platelets | All | 1.14 | 1.01 | 1.27 | 437 | 0.03 |
| | All, Adjusted | 1.16 | 1.04 | 1.29 | 437 | 0.01 |
| | PBO | 1.2 | 1 | 1.45 | 142 | 0.05 |
| | PBO, Adjusted | 1.2 | 1 | 1.44 | 142 | 0.05 |
| log(WBC) | All | 0.65 | 0.58 | 0.74 | 420 | 0 |
| | All, Adjusted | 0.78 | 0.68 | 0.9 | 420 | 0 |
| | PBO | 0.55 | 0.44 | 0.69 | 140 | 0 |
| | PBO, Adjusted | 0.69 | 0.54 | 0.89 | 140 | 0 |

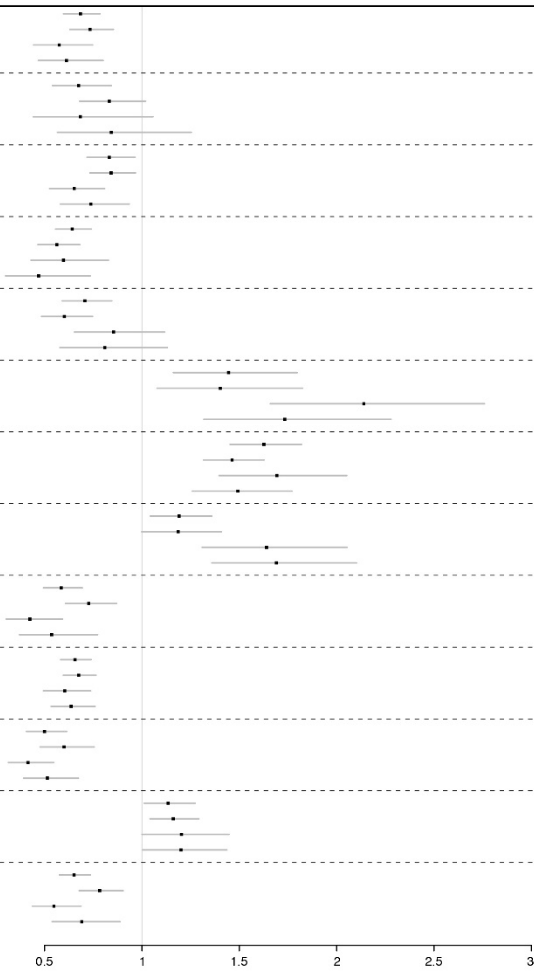

*Appendix Figure S3:* **Predictive forest (tertiles and continuous analysis) plots**

(A) Ordinal scale and (B) time to hospital discharge.

**A**

**Ordinal Scale Day 28; Predictive, Scaled, Adjusted**

| Biomarker | Group | Odds Ratio | Lower | Upper | N | P–Value, Int. |
|---|---|---|---|---|---|---|
| CRP | 0 – 33% | 0.74 | 0.34 | 1.62 | | |
| CRP | 33 – 67% | 1.32 | 0.62 | 2.84 | 382 | 0.31 |
| CRP | 67 – 100% | 0.73 | 0.36 | 1.48 | | |
| CRP | All Subjects | 0.89 | 0.58 | 1.37 | | |
| log(D–Dimer) | 0 – 33% | 0.48 | 0.15 | 1.54 | | |
| log(D–Dimer) | 33 – 67% | 0.33 | 0.11 | 1.01 | 197 | 0.53 |
| log(D–Dimer) | 67 – 100% | 0.97 | 0.32 | 2.98 | | |
| log(D–Dimer) | All Subjects | 0.56 | 0.29 | 1.05 | | |
| log(Ferritin) | 0 – 33% | 1.33 | 0.62 | 2.92 | | |
| log(Ferritin) | 33 – 67% | 0.83 | 0.38 | 1.79 | 364 | 0.02 |
| log(Ferritin) | 67 – 100% | 0.64 | 0.31 | 1.31 | | |
| log(Ferritin) | All Subjects | 0.92 | 0.59 | 1.43 | | |
| log(IL6) | 0 – 33% | 0.78 | 0.32 | 1.94 | | |
| log(IL6) | 33 – 67% | 0.57 | 0.25 | 1.29 | 332 | 0.74 |
| log(IL6) | 67 – 100% | 0.78 | 0.36 | 1.69 | | |
| log(IL6) | All Subjects | 0.73 | 0.46 | 1.18 | | |
| LDH | 0 – 33% | 0.42 | 0.18 | 0.95 | | |
| LDH | 33 – 67% | 1.13 | 0.5 | 2.65 | 364 | 0.27 |
| LDH | 67 – 100% | 0.75 | 0.36 | 1.58 | | |
| LDH | All Subjects | 0.71 | 0.45 | 1.11 | | |
| log(Lymphocytes) | 0 – 33% | 0.84 | 0.44 | 1.61 | | |
| log(Lymphocytes) | 33 – 67% | 1.03 | 0.49 | 2.2 | 426 | 0.39 |
| log(Lymphocytes) | 67 – 100% | 0.8 | 0.37 | 1.77 | | |
| log(Lymphocytes) | All Subjects | 0.9 | 0.6 | 1.37 | | |
| % Lymphocytes | 0 – 33% | 0.61 | 0.31 | 1.19 | | |
| % Lymphocytes | 33 – 67% | 0.9 | 0.43 | 1.88 | 401 | 0.81 |
| % Lymphocytes | 67 – 100% | 0.87 | 0.36 | 2.2 | | |
| % Lymphocytes | All Subjects | 0.76 | 0.48 | 1.2 | | |
| % Monocytes | 0 – 33% | 0.64 | 0.32 | 1.26 | | |
| % Monocytes | 33 – 67% | 0.67 | 0.31 | 1.45 | 400 | 0.11 |
| % Monocytes | 67 – 100% | 1.06 | 0.48 | 2.4 | | |
| % Monocytes | All Subjects | 0.79 | 0.51 | 1.23 | | |
| Neutrophils | 0 – 33% | 1.23 | 0.57 | 2.72 | | |
| Neutrophils | 33 – 67% | 0.4 | 0.19 | 0.83 | 430 | 0.88 |
| Neutrophils | 67 – 100% | 1.12 | 0.6 | 2.12 | | |
| Neutrophils | All Subjects | 0.83 | 0.55 | 1.25 | | |
| % Neutrophils | 0 – 33% | 1.1 | 0.5 | 2.46 | | |
| % Neutrophils | 33 – 67% | 0.69 | 0.32 | 1.53 | 402 | 0.37 |
| % Neutrophils | 67 – 100% | 0.65 | 0.33 | 1.28 | | |
| % Neutrophils | All Subjects | 0.79 | 0.51 | 1.23 | | |
| log(%Neut/%Lym) | 0 – 33% | 1.01 | 0.42 | 2.55 | | |
| log(%Neut/%Lym) | 33 – 67% | 0.85 | 0.41 | 1.8 | 397 | 0.78 |
| log(%Neut/%Lym) | 67 – 100% | 0.61 | 0.31 | 1.19 | | |
| log(%Neut/%Lym) | All Subjects | 0.81 | 0.52 | 1.25 | | |
| Platelets | 0 – 33% | 0.6 | 0.3 | 1.18 | | |
| Platelets | 33 – 67% | 0.65 | 0.31 | 1.36 | 437 | 0.11 |
| Platelets | 67 – 100% | 1.44 | 0.71 | 2.95 | | |
| Platelets | All Subjects | 0.82 | 0.55 | 1.23 | | |
| log(WBC) | 0 – 33% | 0.74 | 0.34 | 1.6 | | |
| log(WBC) | 33 – 67% | 0.54 | 0.26 | 1.14 | 420 | 0.77 |
| log(WBC) | 67 – 100% | 0.87 | 0.46 | 1.67 | | |
| log(WBC) | All Subjects | 0.72 | 0.47 | 1.09 | | |

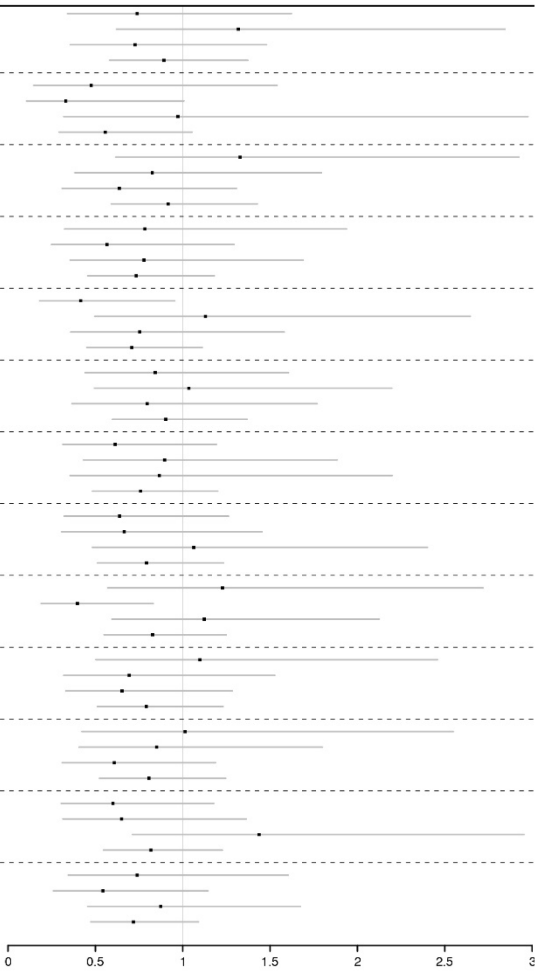

**B**

##### Time to Hospital Discharge; Fine Gray; Predictive, Scaled, Adjusted

| Biomarker | Group | Hazard Ratio | Lower | Upper | N | P–Value, Int. |
|---|---|---|---|---|---|---|
| CRP | 0 – 33% | 1.13 | 0.72 | 1.76 | | |
| CRP | 33 – 67% | 1.11 | 0.64 | 1.94 | 382 | 0.08 |
| CRP | 67 – 100% | 1.8 | 1.01 | 3.21 | | |
| CRP | All Subjects | 1.34 | 0.99 | 1.82 | | |
| log(D–Dimer) | 0 – 33% | 2.1 | 0.99 | 4.45 | | |
| log(D–Dimer) | 33 – 67% | 1.38 | 0.71 | 2.69 | 197 | 0.67 |
| log(D–Dimer) | 67 – 100% | 1.19 | 0.58 | 2.44 | | |
| log(D–Dimer) | All Subjects | 1.49 | 1 | 2.22 | | |
| log(Ferritin) | 0 – 33% | 1.33 | 0.82 | 2.18 | | |
| log(Ferritin) | 33 – 67% | 1.33 | 0.81 | 2.18 | 364 | 0.09 |
| log(Ferritin) | 67 – 100% | 1.78 | 0.97 | 3.26 | | |
| log(Ferritin) | All Subjects | 1.5 | 1.1 | 2.05 | | |
| log(IL6) | 0 – 33% | 1.17 | 0.72 | 1.9 | | |
| log(IL6) | 33 – 67% | 2.74 | 1.49 | 5.03 | 332 | 0.9 |
| log(IL6) | 67 – 100% | 1.93 | 0.86 | 4.35 | | |
| log(IL6) | All Subjects | 1.68 | 1.17 | 2.42 | | |
| LDH | 0 – 33% | 2.29 | 1.46 | 3.6 | | |
| LDH | 33 – 67% | 1.03 | 0.64 | 1.64 | 364 | 0.09 |
| LDH | 67 – 100% | 1.32 | 0.69 | 2.53 | | |
| LDH | All Subjects | 1.47 | 1.08 | 2.01 | | |
| log(Lymphocytes) | 0 – 33% | 1.34 | 0.76 | 2.38 | | |
| log(Lymphocytes) | 33 – 67% | 1.02 | 0.67 | 1.56 | 426 | 0.2 |
| log(Lymphocytes) | 67 – 100% | 1.75 | 1.1 | 2.8 | | |
| log(Lymphocytes) | All Subjects | 1.4 | 1.04 | 1.88 | | |
| % Lymphocytes | 0 – 33% | 2.23 | 1.15 | 4.31 | | |
| % Lymphocytes | 33 – 67% | 1.44 | 0.84 | 2.48 | 401 | 0.79 |
| % Lymphocytes | 67 – 100% | 1.29 | 0.86 | 1.94 | | |
| % Lymphocytes | All Subjects | 1.59 | 1.17 | 2.15 | | |
| % Monocytes | 0 – 33% | 2.11 | 1.06 | 4.18 | | |
| % Monocytes | 33 – 67% | 1.63 | 1.02 | 2.6 | 400 | 0 |
| % Monocytes | 67 – 100% | 1.24 | 0.79 | 1.94 | | |
| % Monocytes | All Subjects | 1.56 | 1.15 | 2.1 | | |
| Neutrophils | 0 – 33% | 1.02 | 0.68 | 1.55 | | |
| Neutrophils | 33 – 67% | 1.86 | 1.08 | 3.2 | 430 | 0.07 |
| Neutrophils | 67 – 100% | 1.57 | 0.86 | 2.86 | | |
| Neutrophils | All Subjects | 1.51 | 1.09 | 2.08 | | |
| % Neutrophils | 0 – 33% | 1.24 | 0.82 | 1.9 | | |
| % Neutrophils | 33 – 67% | 1.52 | 0.89 | 2.61 | 402 | 0.57 |
| % Neutrophils | 67 – 100% | 1.97 | 1.06 | 3.66 | | |
| % Neutrophils | All Subjects | 1.58 | 1.17 | 2.13 | | |
| log(%Neut/%Lym) | 0 – 33% | 1.28 | 0.85 | 1.93 | | |
| log(%Neut/%Lym) | 33 – 67% | 1.47 | 0.83 | 2.62 | 397 | 0.26 |
| log(%Neut/%Lym) | 67 – 100% | 2.16 | 1.14 | 4.09 | | |
| log(%Neut/%Lym) | All Subjects | 1.61 | 1.16 | 2.24 | | |
| Platelets | 0 – 33% | 1.87 | 1.05 | 3.33 | | |
| Platelets | 33 – 67% | 1.39 | 0.88 | 2.2 | 437 | 0.48 |
| Platelets | 67 – 100% | 1.05 | 0.67 | 1.62 | | |
| Platelets | All Subjects | 1.4 | 1.05 | 1.85 | | |
| log(WBC) | 0 – 33% | 1.28 | 0.85 | 1.94 | | |
| log(WBC) | 33 – 67% | 1.65 | 0.98 | 2.77 | 420 | 0.18 |
| log(WBC) | 67 – 100% | 1.88 | 1 | 3.55 | | |
| log(WBC) | All Subjects | 1.58 | 1.17 | 2.15 | | |

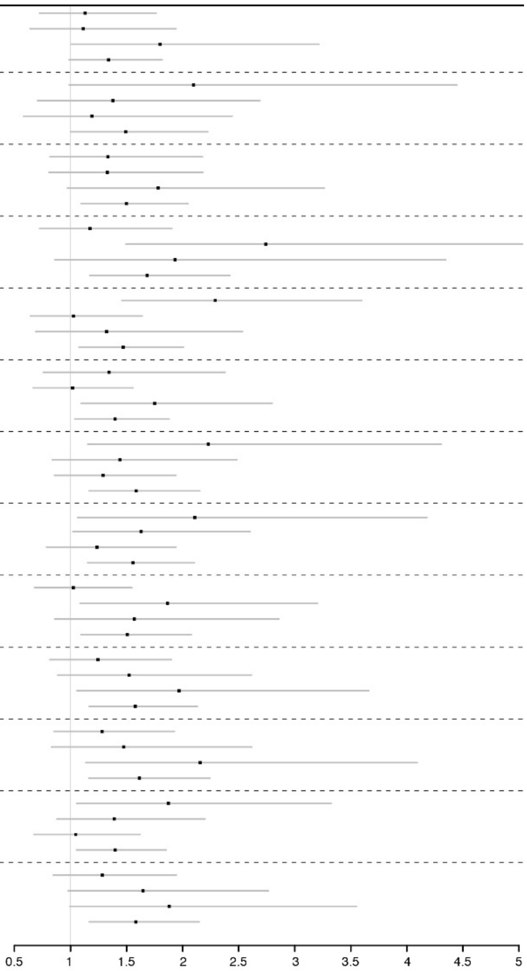

***Appendix Figure S4:***

(A) Ferritin as a predictive biomarker for death by day 28 in combined analysis of COVACTA and MARIPOSA. (B) Baseline ferritin levels in MARIPOSA and COVACTA. (C) Love plot showing the standardised mean difference using matched subjects, unadjusted, and ATT weights. (D) Descriptive plot of subgroups of patients receiving or not receiving mechanical ventilation in MARIPOSA and COVACTA.

**A**

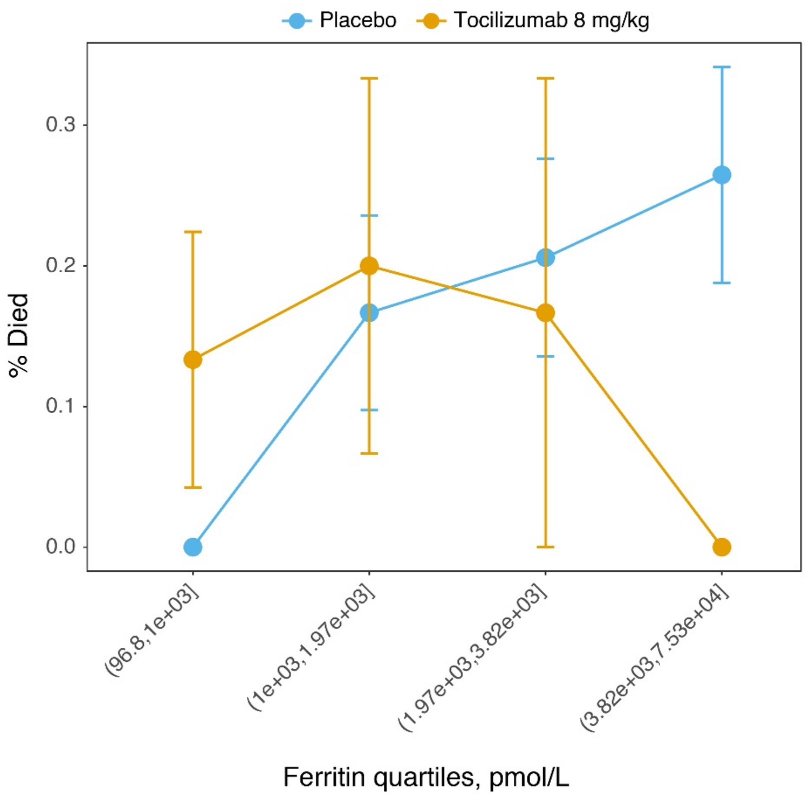

**B**

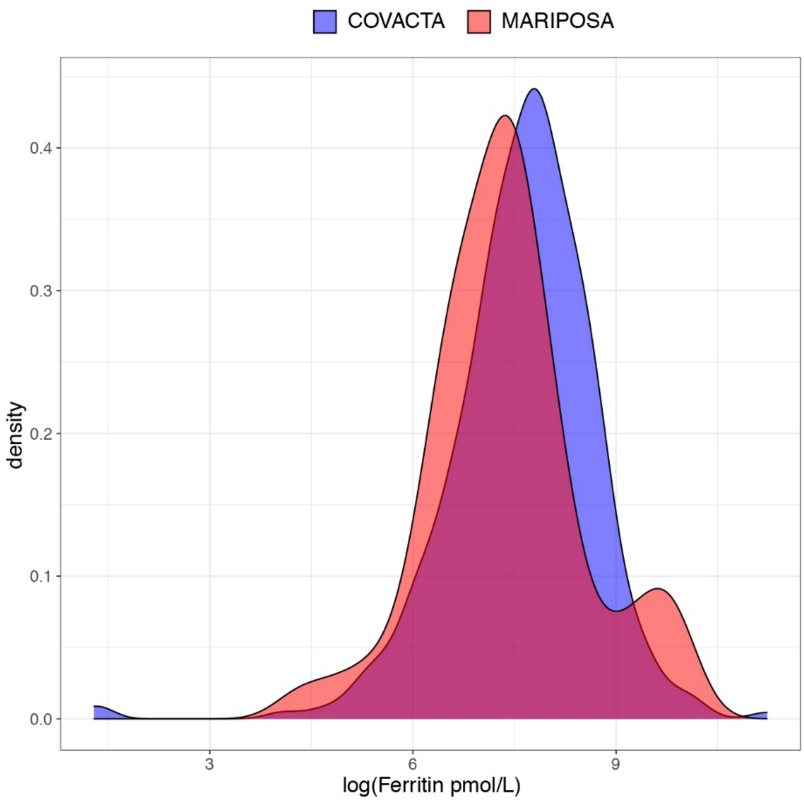
COVACTA
MARIPOSA
density
0.0
0.1
0.2
0.3
0.4
3
6
9
log(Ferritin pmol/L)

**C**

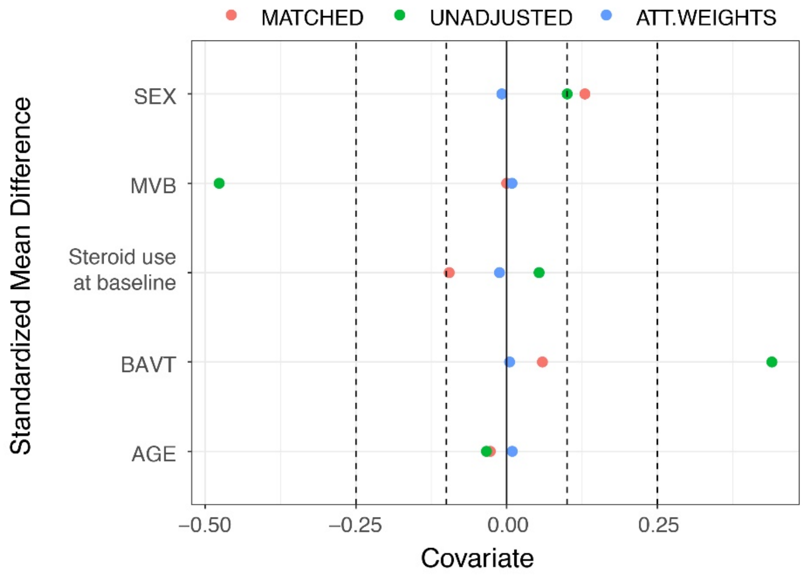
MATCHED
UNADJUSTED
ATT.WEIGHTS
Standardized Mean Difference
SEX
MVB
Steroid use
at baseline
BAVT
AGE
−0.50
−0.25
0.00
0.25
Covariate

**D**

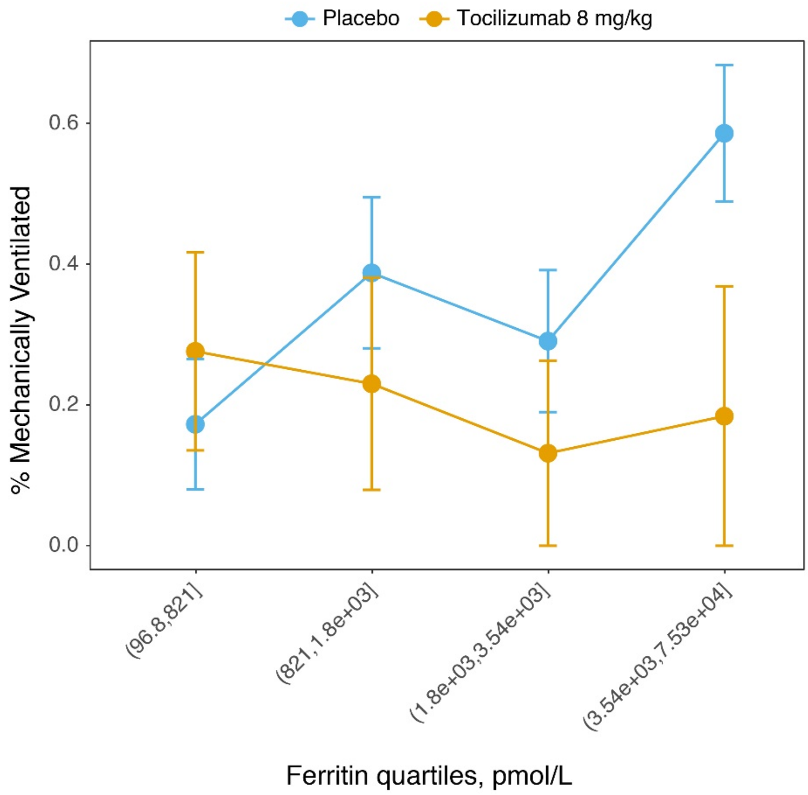


(A, D) Data are shown as mean ± standard error based on 124 patients from the placebo arm of COVACTA and 67 patients from the tocilizumab 4-mg/kg and 8-mg/kg arms of MARIPOSA.

BAVT, baseline antiviral treatment; MVB, mechanical ventilation at baseline; PBO=placebo.

***Appendix Figure S5:*** **Day 28 outcomes by baseline IL-6 tertiles (mITT population)**

(A) Ordinal scale category and (B) death.

**A**

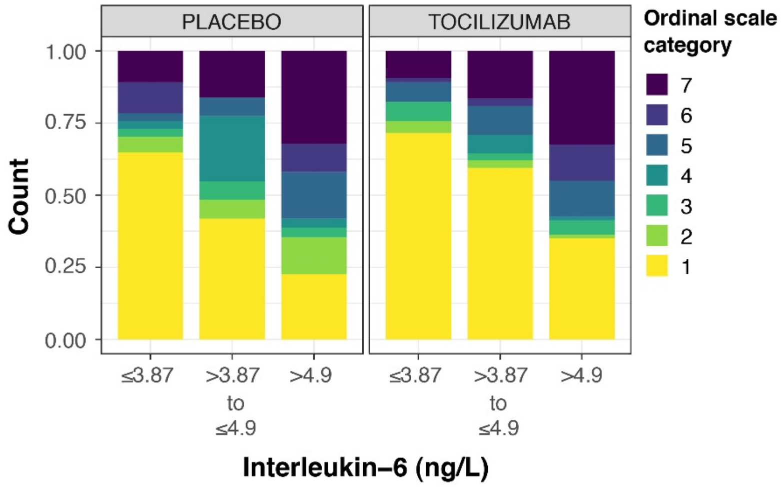

**B**

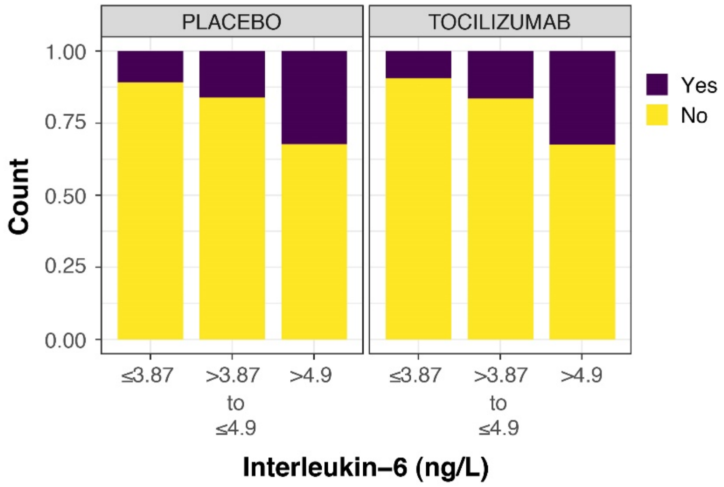


Seven-category ordinal scale: 1, discharged or ready for discharge; 2, non–ICU hospital ward, not requiring supplemental oxygen; 3, non–ICU hospital ward, requiring supplemental oxygen; 4, ICU or non–ICU hospital ward, requiring non-invasive ventilation or high-flow oxygen; 5, ICU, requiring intubation and mechanical ventilation; 6, ICU, requiring extracorporeal membrane oxygenation or mechanical ventilation and additional organ support; 7, death.

IL-6 values are log transformed.

ICU=intensive care unit; IL-6=interleukin-6.

***Appendix Figure S6:***

Unmodelled data by ferritin quartiles (A) in all-comers (N=437) and (B) in the subgroup of patients requiring positive pressure or mechanical ventilation at baseline (ordinal scale categories 4 and 5) (n=157). (C) Baseline ordinal scale versus baseline ferritin levels.

**A**

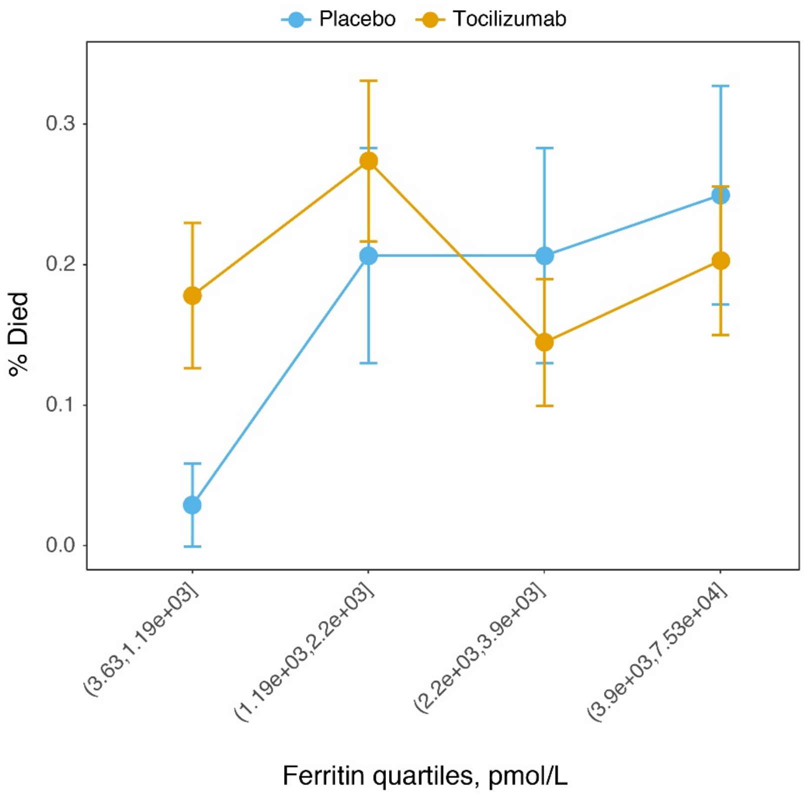

**B**

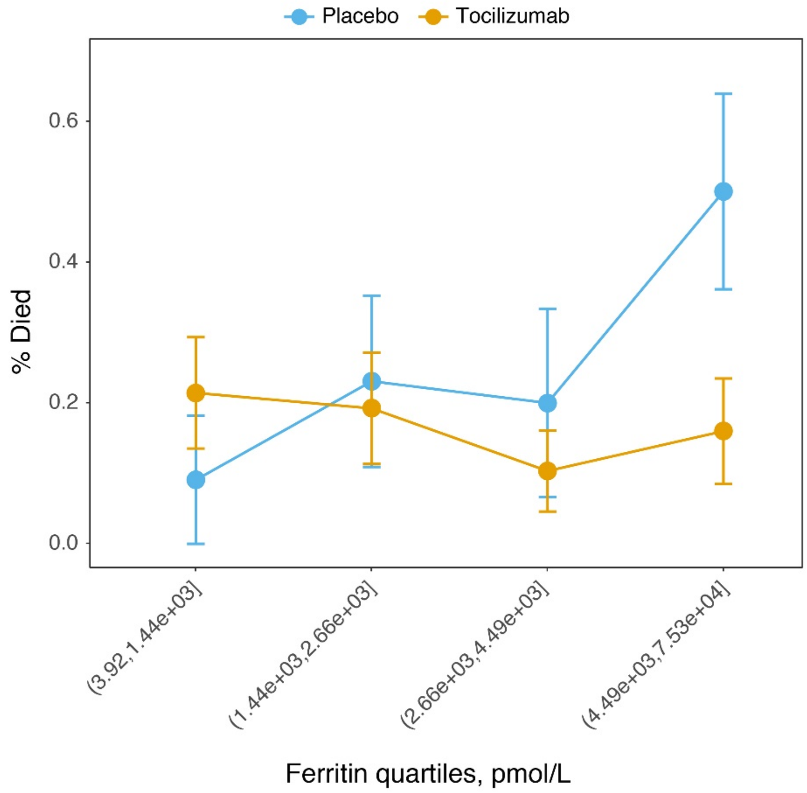
Placebo
Tocilizumab
% Died
0.6
0.4
0.2
0.0
(3.92,1.44e+03]
(1.44e+03,2.66e+03]
(2.66e+03,4.49e+03]
(4.49e+03,7.53e+04]
Ferritin quartiles, pmol/L

**C**

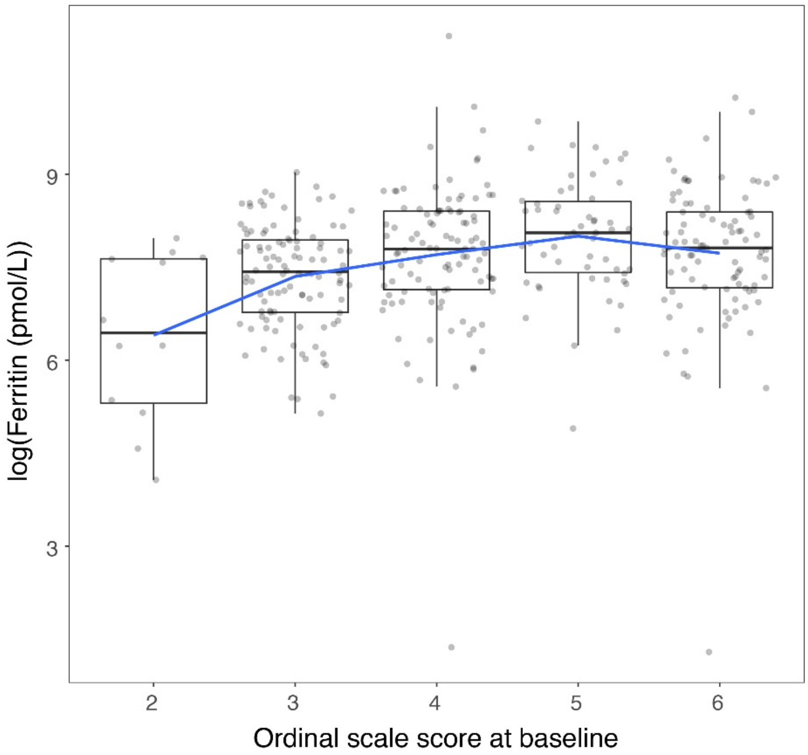


(A, B) Data are shown as descriptive mean ± SE. (C) The blue line shows Loess smoothing.

SE=standard error.

***Appendix Figure S7:***

(A) Prognostic models and (B) predictive models for clinical outcomes by biomarkers and by ferritin tertiles (C, ordinal scale score; D, mechanical ventilation status; E, mortality; F, hospital discharge) in the subgroup of patients requiring positive pressure or mechanical ventilation at baseline (ordinal scale categories 4 and 5) (n=157).

**A**

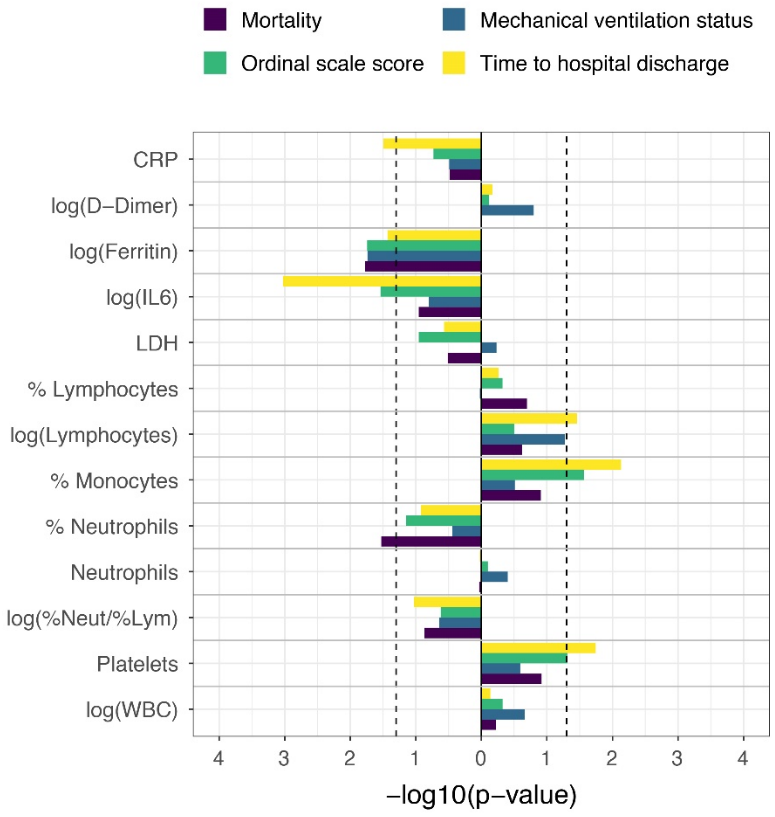

**B**

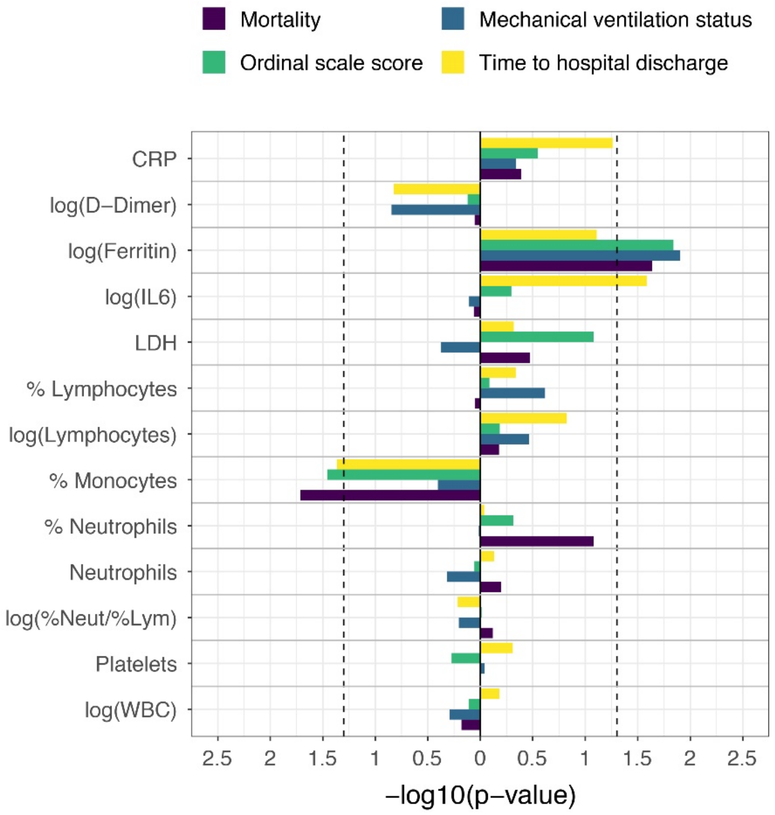

Mortality
Mechanical ventilation status
Ordinal scale score
Time to hospital discharge
CRP
log(D−Dimer)
log(Ferritin)
log(IL6)
LDH
% Lymphocytes
log(Lymphocytes)
% Monocytes
% Neutrophils
Neutrophils
log(%Neut/%Lym)
Platelets
log(WBC)
2.5 2 1.5 1 0.5 0 0.5 1 1.5 2 2.5
−log10(p−value)

C

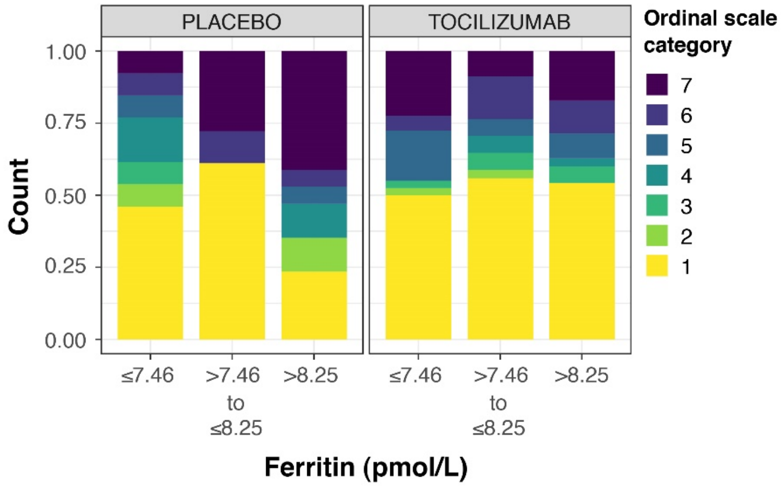
PLACEBO
TOCILIZUMAB
Ordinal scale category
7
6
5
4
3
2
1
Count
1.00
0.75
0.50
0.25
0.00
≤7.46
>7.46 to ≤8.25
>8.25
Ferritin (pmol/L)

**D**

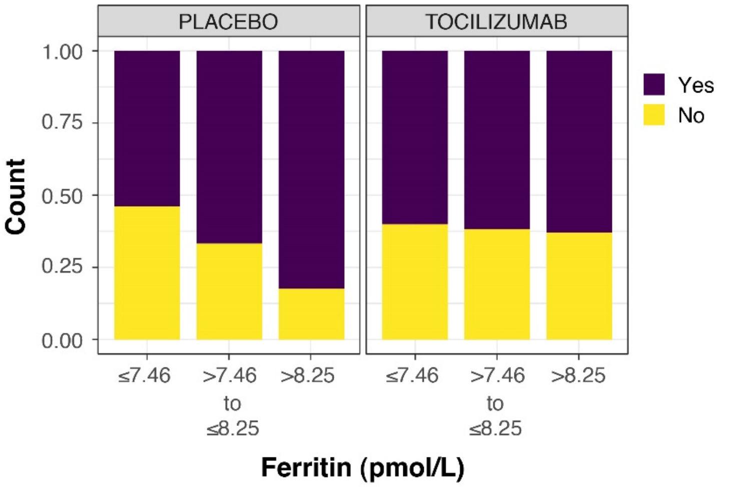
PLACEBO
TOCILIZUMAB
1.00
0.75
0.50
0.25
0.00
Count
≤7.46
>7.46 to ≤8.25
>8.25
Ferritin (pmol/L)
Yes
No

**E**

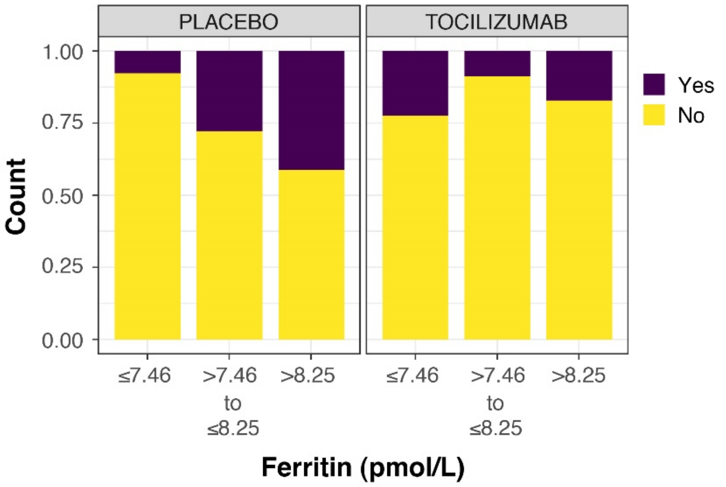
PLACEBO
TOCILIZUMAB
1.00
0.75
0.50
0.25
0.00
Count
≤7.46
>7.46 to ≤8.25
>8.25
Ferritin (pmol/L)
Yes
No

**F**

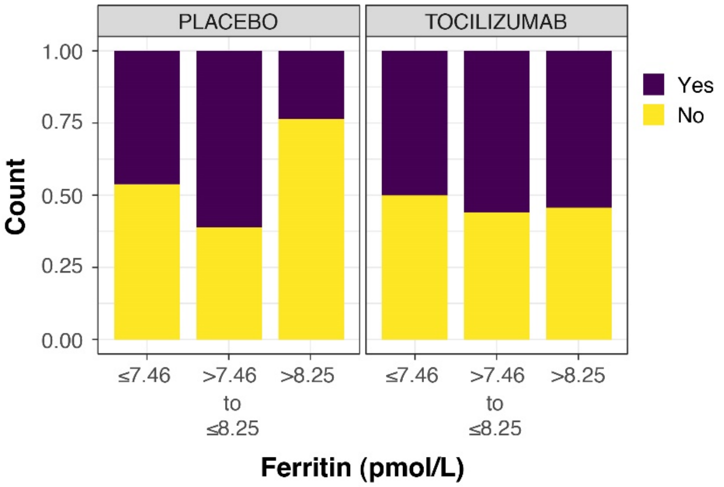


Seven-category ordinal scale: 1, discharged or ready for discharge; 2, non–ICU hospital ward, not requiring supplemental oxygen; 3, non–ICU hospital ward, requiring supplemental oxygen; 4, ICU or non–ICU hospital ward, requiring non-invasive ventilation or high-flow oxygen; 5, ICU, requiring intubation and mechanical ventilation; 6, ICU, requiring extracorporeal membrane oxygenation or mechanical ventilation and additional organ support; 7, death.

ICU=intensive care unit.

*Appendix Figure S8:* **Cumulative probability of time to discharge and death by day 28 according to baseline IL-6 concentration in patients requiring positive pressure or mechanical ventilation at baseline (ordinal scale categories 4 and 5) (n=157)**

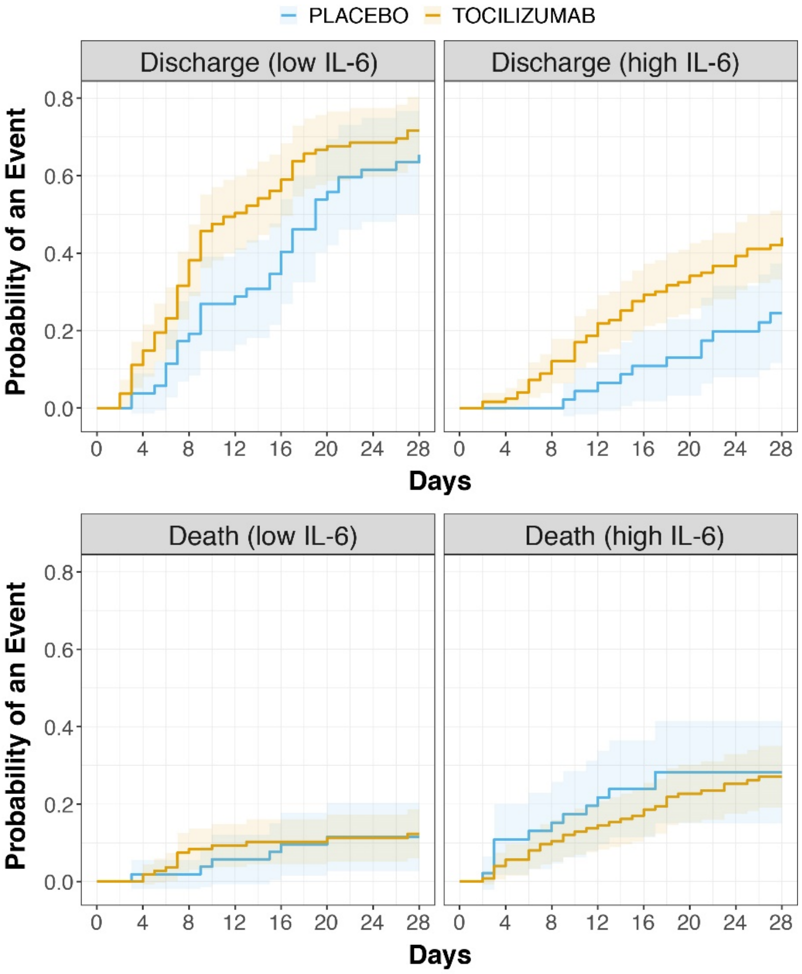


IL-6 values are log transformed. A cut-off IL-6 value of 80 ng/L was chosen for visualisation purposes only (C) based on the median (IQR) value of 85·8 (35·6–188·0) ng/mL.

IL-6=interleukin-6; IQR=interquartile range.
